# A unified model for staging amyloid and tau pathology in Alzheimer’s disease

**DOI:** 10.64898/2026.03.30.26349752

**Authors:** Tom W. Earnest, Braden Y. Yang, Ahmad Chowdhury, Sung Min Ha, Abdalla Bani, Soo-Jong Kim, Arash Nazeri, John C. Morris, Tammie L. S. Benzinger, Brian A. Gordon, the Alzheimer’s Disease Neuroimaging Initiative, The HABS-HD Study Team, Aristeidis Sotiras

**Affiliations:** Mallinckrodt Institute of Radiology, Washington University School of Medicine in St. Louis; St. Louis, MO, USA; Department of Radiology, Perelman School of Medicine, University of Pennsylvania; Philadelphia, PA, USA; Center for AI and Data Science for Integrated Diagnostics (AI2D), Perelman School of Medicine, University of Pennsylvania; Philadelphia, PA, USA; Department of Neurology, Washington University School of Medicine in St. Louis; St. Louis, St. Louis, MO, USA; Institute for Informatics, Data Science and Biostatistics, Washington University School of Medicine in St. Louis; St. Louis, MO, USA

## Abstract

Biological staging models are a key tool for assessing the severity of Alzheimer’s disease (AD), supporting personalized medicine and playing a critical role in clinical trial design. Recently, researchers have leveraged positron emission tomography (PET) to inform data-driven staging models of brain pathology related to AD. However, most approaches have focused on staging either amyloid or tau progressions separately, while both pathologies constitute defining factors of AD. Here, we aimed to derive a data-driven staging model which encompasses the spatial spread of both amyloid and tau. We assembled a large sample (n=3,293) of individuals with both amyloid and tau PET imaging stemming from 8 neuroimaging studies of AD and aging. We applied unsupervised machine learning to estimate brain areas which showed coordinated pathological accumulation across our sample, and we used these regions to inform a data-driven model for staging amyloid and tau. The resulting six stage model showed two stages of amyloid progression followed by four stages of tau spread, which were associated with cross-sectional and longitudinal assessments of cognitive decline. Comparison of our biological staging model with clinical disease stages recommended by the Alzheimer’s Association showed evidence of heterogenous symptom profiles. Replication of results in holdout data demonstrated the generalizability and prognostic value of our staging model. Together, these findings establish a comprehensive and rigorously validated biological staging model that jointly characterizes amyloid and tau progression, advances beyond global or anatomically predefined summaries, and provides a scalable framework for studying disease heterogeneity and progression in AD.

**One Sentence Summary:** Using PET imaging from a large sample of individuals (n=3,293), we derive a data-driven model for staging amyloid and tau pathology.

## INTRODUCTION

Alzheimer’s disease (AD) is a neurodegenerative disorder and the leading cause of dementia worldwide. Biologically, AD is defined by the formation of two core proteinopathies, namely amyloid plaques and tau neurofibrillary tangles (*1*). Amyloid pathology typically forms 10-20 years before the onset of overt cognitive symptoms, constituting a lengthy preclinical phase. Tau deposition, closely related to neuronal death, appears more proximally to cognitive impairment. Treatments which target these pathological processes are currently limited, with the only approved drugs being monoclonal antibodies which target amyloid plaques (*2, 3*). Crucially, effective administration of these drugs requires early intervention in the disease course before neurodegenerative processes have caused irrevocable damage. Moreover, the development of novel interventions will require methods for stratifying individuals based on current disease severity and prognostic risk of decline (*4*). As such, disease staging models, which provide biomarker-based assessments of disease severity, are a key tool for administering and developing AD treatments.

While disease staging models for AD were originally based on neuropathological studies (*5, 6*), the advent of positron emission tomography (PET) ligands specific to amyloid and tau has allowed *in vivo* disease staging. Namely, an individual’s PET data can be compared against a model which outlines the spatial trajectories of proteinopathies with advancing disease. For amyloid, however, modern PET-based analyses typically rely on a dichotomous positive/negative assessment of amyloid pathology, ignoring the spatial progression which may be informative of prognosis (*1, 7*). Alternatively, tau staging has received more focus, with several PET studies applying the Braak system (*8–11*). Still, there are methodological considerations which may complicate the translation of neuropathological findings to PET imaging. Staging models derived from autopsy are conducted in restricted samples, much smaller and less diverse than what is tractable with PET. Additionally, neuropathological studies are informed by sparse sampling of brain regions, which may miss larger brain-spanning patterns observable with PET.

Given these limitations, some approaches have instead applied data-driven methods to derive staging systems directly from PET imaging. These studies typically infer disease progressions from cross-sectional distributions of regional pathology. This approach was first applied for amyloid, with resulting models exhibiting three (*12*) or four stages (*7, 13*). More recent studies have also applied similar methods for tau-PET (*14*), including our own work (*15*). Other approaches have explicitly aimed to capture heterogenous disease progressions, modelling amyloid and tau subtypes with unique spatiotemporal progressions (*16–20*).

While data-driven staging studies to date have been highly informative for understanding AD pathological progression, they suffer from three key limitations. First, most designs are reliant on conventional anatomic atlases and handpicked regions of interest (ROIs). This approach restricts disease stages to arbitrary spatial patterns not informed by AD pathology. These *a priori* regional boundaries discard the rich spatial information present in PET images. Second, prior approaches to staging have determined the number of disease stages using methods which are unrelated to the underlying biological complexity. For example, the number of stages may be chosen ad hoc (*7, 13*) or proportional to the number of biomarkers included (*16, 17*). Ideally, data-driven methods should be used to identify stages which capture biologically meaningful changes in biomarker presentations. Third, most studies have focused on only staging amyloid or tau independently, not considering the joint progression of both pathologies. Recent consensus research guidelines for AD have stressed the importance of biological staging models that incorporate both amyloid and tau and have outlined a staging framework based on amyloid PET and tau PET (*1*). However, this framework is primarily conceptual and leaves considerable ambiguity regarding implementation. There is a critical gap between theoretical staging frameworks and their operational implementation using *in vivo* PET imaging data. Novel approaches should delineate the spatiotemporal progressions of amyloid and tau in the same sample, applying rigorous, data-driven methods to both pathologies to derive a reproducible and comprehensive biological staging scheme for AD.

In this study, we aimed to address these gaps by developing a unified, data-driven staging model for amyloid and tau pathology (Figure 1). We assembled a large sample of individuals with both amyloid-PET and tau-PET imaging to model the evolution of both core proteinopathies of AD in the same individuals. Rather than relying on a predefined anatomical atlas, we pursued a voxel-based analysis to uncover high-dimensional spatial patterns of pathological accumulation. To identify the brain areas key to disease progression, we applied non-negative matrix factorization (NMF), an unsupervised machine learning method that detects areas of coordinated signal covariance. We supplied the NMF-estimated hubs of amyloid and tau deposition to a staging algorithm to empirically determine stages of spatiotemporal disease progression. We finally used cross-sectional and longitudinal assessments to demonstrate the prognostic value of our staging model. Throughout our analysis, we used independent data for replication of our model development and validation of findings. Our results demonstrate a comprehensive model for assessing the core pathologies of AD, shedding light on its biological etiology and presenting a tool for individualized disease staging.

**Figure 1.**
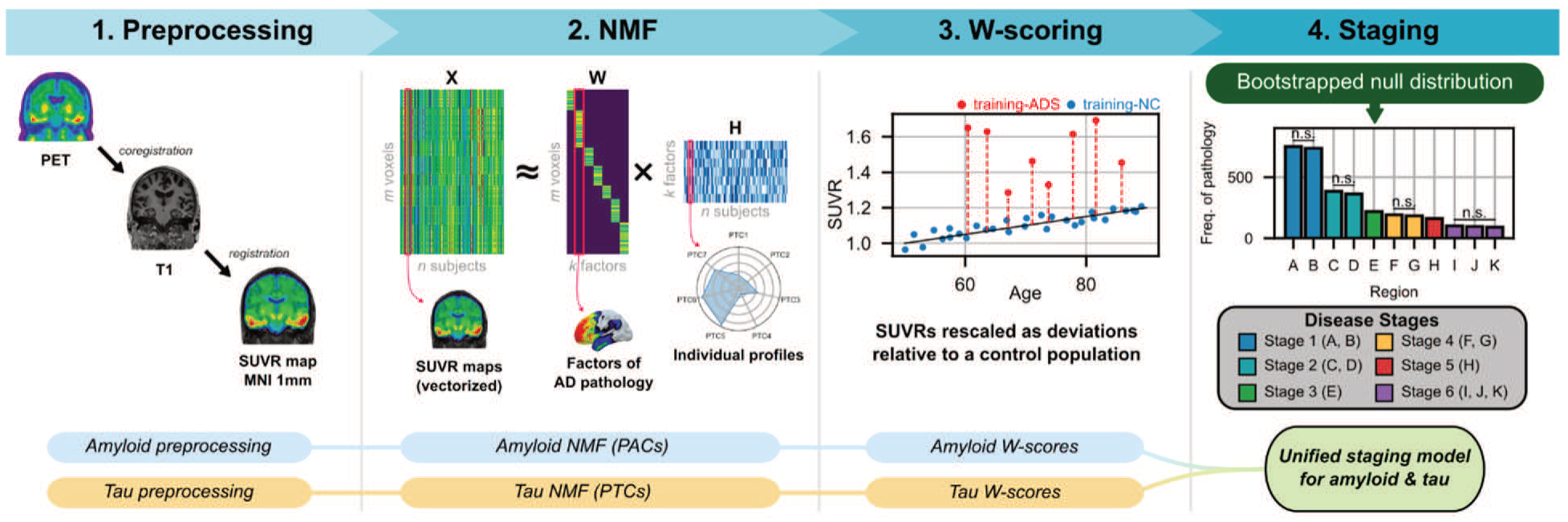
Overview of methods. Amyloid and tau images start under separate processing streams. Each modality is preprocessed using an MRI-based pipeline to generate SUVR maps in standard space (1. Preprocessing). Registered amyloid and tau PET images are then supplied to NMF to estimate factors capturing areas of coordinated pathology development (PACs/PTCs) (2. NMF). Individual uptakes within PACs and PTCs are rescaled to normative deviations (W-scores) by modeling uptake in the normative control group (3. W-scoring). W-scores for PACs and PTCs are integrated in an algorithm for automated determination of disease stages (4. Staging).

## RESULTS

We incorporated 3,293 individuals from 8 different datasets (Table 1, Supplementary Figure 1). Included individuals were scanned using four different tracer pairs: [^18^F]-florbetapir (FBP) and [^18^F]-flortaucipir (FTP), [^18^F]-florbetaben (FBB) and FTP, FBB and [^18^F]-PI-2620 (PI2620), or [^11^C]-Pittsburgh Compound B (PIB) and FTP. We established a fixed-tracer training set consisting of FBP/FTP scans (n=1,387) and a mixed tracer validation set including heterogeneous tracer pairs (n=1,906; FBP/FTP: n=323; FBB/P26: n=1,097; PIB/FTP: n=486). Both training and validation sets contained individuals on the Alzheimer’s Disease spectrum (ADS, positive for amyloid pathology; training-ADS: n=894; validation-ADS: n=754) and cognitively unimpaired individuals without AD pathology who served as normative controls (NC; training-NC: n=493, validation-NC: n=1,152).

**Table 1.** Descriptive statistics of the sample. Continuous variables are represented as mean (standard deviation) and compared across groups using a two-sample t-test. Categorical variables are presented as count (percentage%) and are compared across groups using Chi-squared tests. The “N” column shows the number of individuals with non-missing observations. ADS=Alzheimer’s Disease Spectrum; BMI=body mass index; APOEE4=apolipoprotein E (E4 carriership); CDR=Clinical Dementia Rating (global score); FBB=florbetaben; FBP=florbetapir; FTP=flortaucipir; P26=PI-2620; PIB=Pittsburgh Compount B; MMSE=Mini Mental State Examination (total score); SUVR=standardized uptake value ratio.

| Characteristic | N | Training N = 1,387 | Validation N = 1,906 | p-value |
| --- | --- | --- | --- | --- |
| Disease | 3,293 |  |  | <0.001 |
| ADS |  | 894 (64%) | 754 (40%) |  |
| NC |  | 493 (36%) | 1,152 (60%) |  |
| Age | 3,293 | 72.11 (7.25) | 69.17 (8.87) | <0.001 |
| Sex | 3,293 |  |  | 0.049 |
| M |  | 591 (43%) | 747 (39%) |  |
| F |  | 796 (57%) | 1,159 (61%) |  |
| Race | 3,263 |  |  | <0.001 |
| White |  | 1,205 (88%) | 1,463 (77%) |  |
| Black |  | 100 (7.3%) | 396 (21%) |  |
| Asian |  | 37 (2.7%) | 25 (1.3%) |  |
| Other |  | 20 (1.5%) | 17 (0.9%) |  |
| Hispanic | 3,142 | 42 (3.4%) | 311 (16%) | <0.001 |
| Education | 3,141 | 16.38 (2.58) | 15.44 (3.05) | <0.001 |
| BMI | 2,225 | 27.63 (5.17) | 29.74 (6.68) | <0.001 |
| CDR | 3,225 |  |  | 0.002 |
| 0.0 |  | 1,181 (85%) | 1,483 (81%) |  |
| 0.5 |  | 157 (11%) | 280 (15%) |  |
| 1.0+ |  | 46 (3.3%) | 78 (4.2%) |  |
| MMSE | 2,910 | 28.44 (2.26) | 28.14 (2.46) | <0.001 |
| APOEE4 | 3,062 |  |  | <0.001 |
| Negative |  | 664 (50%) | 1,161 (67%) |  |
| Positive |  | 666 (50%) | 571 (33%) |  |
| Tracers | 3,293 |  |  | <0.001 |
| FBP/FTP |  | 1,387 (100%) | 0 (0%) |  |
| FBB/FTP |  | 0 (0%) | 323 (17%) |  |
| FBB/P26 |  | 0 (0%) | 1,097 (58%) |  |
| PIB/FTP |  | 0 (0%) | 486 (25%) |  |
| Amyloid SUVR | 3,293 | 1.36 (0.25) | 1.34 (0.34) | 0.024 |
| Tau SUVR | 3,293 | 1.21 (0.20) | 1.17 (0.24) | <0.001 |
| Visits | 3,293 |  |  |  |
| 1 |  | 894 (64%) | 1,345 (71%) |  |
| 2 |  | 423 (30%) | 499 (26%) |  |
| 3 |  | 44 (3.2%) | 55 (2.9%) |  |
| 4 |  | 23 (1.7%) | 7 (0.4%) |  |
| 5 |  | 3 (0.2%) | 0 (0%) |  |
| Followup (y) | 1,054 | 2.31 (1.95) | 2.54 (1.22) | 0.025 |

### Data-driven regions of amyloid and tau accumulation

We first aimed to identify brain regions to inform the staging of amyloid and tau pathology using NMF. NMF is an unsupervised machine learning approach which identifies covariance structures (factors) highlighting areas of coordinated variation across an input dataset. Importantly, the non-negativity constraints of NMF engender factors which are more interpretable than alternative approaches (*21*). Applied to neuroimaging, NMF has been able to extract anatomically meaningful areas of brain variability which shed light on disease and normal aging (*15, 21, 22*). Here, we applied NMF to voxel-level amyloid and tau SUVR maps of training subjects to identify factors which capture the effective spatial resolution of each pathology. Balancing data fit and generalizability, our model selection procedure identified 4 amyloid factors and 7 tau factors after manual removal of white matter and reference tissue components (Supplementary Figure 2, Supplementary Figure 3). We call these factors Patterns of Amyloid Covariance (PACs) and Patterns of Tau Covariance (PTCs) (Figure 2A, B). PACs and PTCs mapped onto identifiable brain regions and were predominantly symmetrical across hemispheres. For amyloid, cortical areas were primarily captured by two factors, one involving the frontal lobe and portions of the cingulate (PAC-Frontal) and another highlighting precuneus and lateral parietal cortices (PAC-Parietal). The remaining amyloid factors involved sensory and motor regions, including the occipital cortex (PAC-Occipital) and areas surrounding the central sulcus (PAC-Sensorimotor). Primary sensory and motor regions were also identified for tau (PTC-Occipital, PTC-Sensorimotor), although other brain areas loaded differentially onto tau factors. The frontal lobe was represented by two factors, one encompassing dorsal and lateral regions (PTC-Frontal) and another capturing medial, orbitofrontal, and insular cortices (PTC-InsularMedialFrontal). Lateralized tau factors were observed in the parietal and temporal lobes (PTC-LeftParietalTemporal, PTC-RightParietalTemporal). A final tau factor covered medial temporal structures (PTC-MedialTemporal).

**Figure 2.**
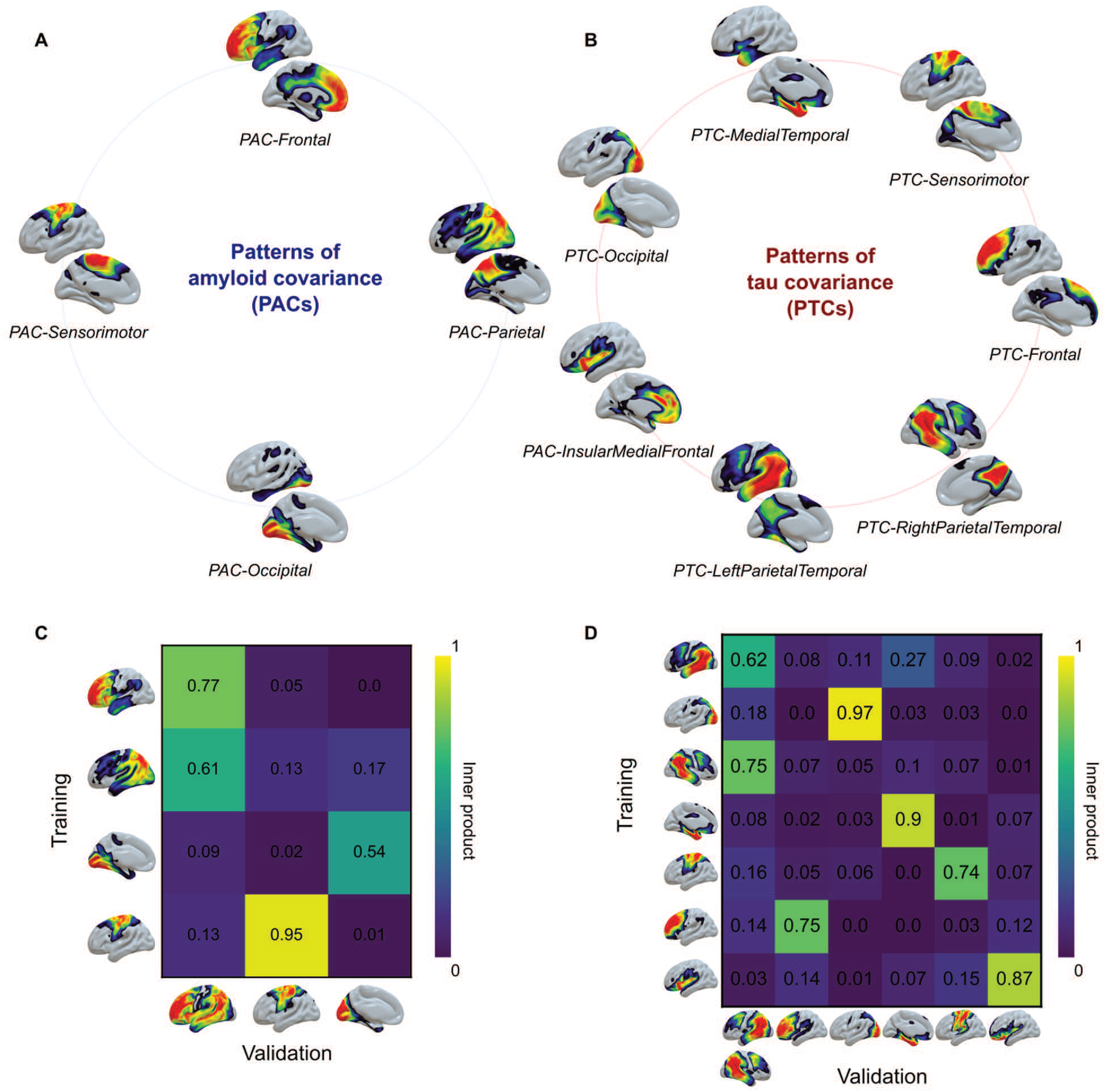
Data-driven factors of amyloid and tau estimated with non-negative matrix factorization (NMF). **(A)** The four selected amyloid factors. **(B)** The seven selected tau factors. **(C)** Heatmap showing similarity (inner product) of matched amyloid factors estimated in training (rows) and validation (columns). **(D)** Same as (C), but for tau.

Using validation data, we retrained amyloid and tau NMF models to evaluate model reproducibility. Despite the validation set including different data sources and PET tracers, we found highly similar factors (Figure 2C, D). Most training factors had a single clear analogue in the validation set (maximum inner product: 0.54-0.95). The largest discrepancies arose when validation factors reflected combinations of training factors (Supplementary Figure 4). PAC-Frontal and PAC-Parietal appeared as a single validation factor, capturing areas typically used for amyloid quantification (*23*) (Supplementary Figure 5). Similarly, PTC-LeftParietalTemporal and PTC-RightParietalTemporal were merged into a single bilateral validation factor. Factors were also largely reproducible in fixed-tracer subsets of the validation data, with differences again due to hierarchical merging of factors (Supplementary Figure 6, Supplementary Figure 7).

### An integrated amyloid and tau PET staging model

We next aimed to determine the temporal progression of pathology across PACs and PTCs. Following conventional approaches (*13, 17*), we used the rates of elevated PET uptake across regions to inform the temporal progression of pathology. To assess regional rates of pathology, we calculated standardized uptake value ratios (SUVRs) within PACs and PTCs for all subjects. We converted PET uptakes of the training-ADS cohort into W-scores (*15, 24*), generating individualized measures of ADS-related pathology as deviations from a healthy distribution (training-NC). We determined positivity for pathology across PACs and PTCs (W>2.5), which indicated a spread of pathology for both amyloid and tau (Figure 3A & B).

**Figure 3.**
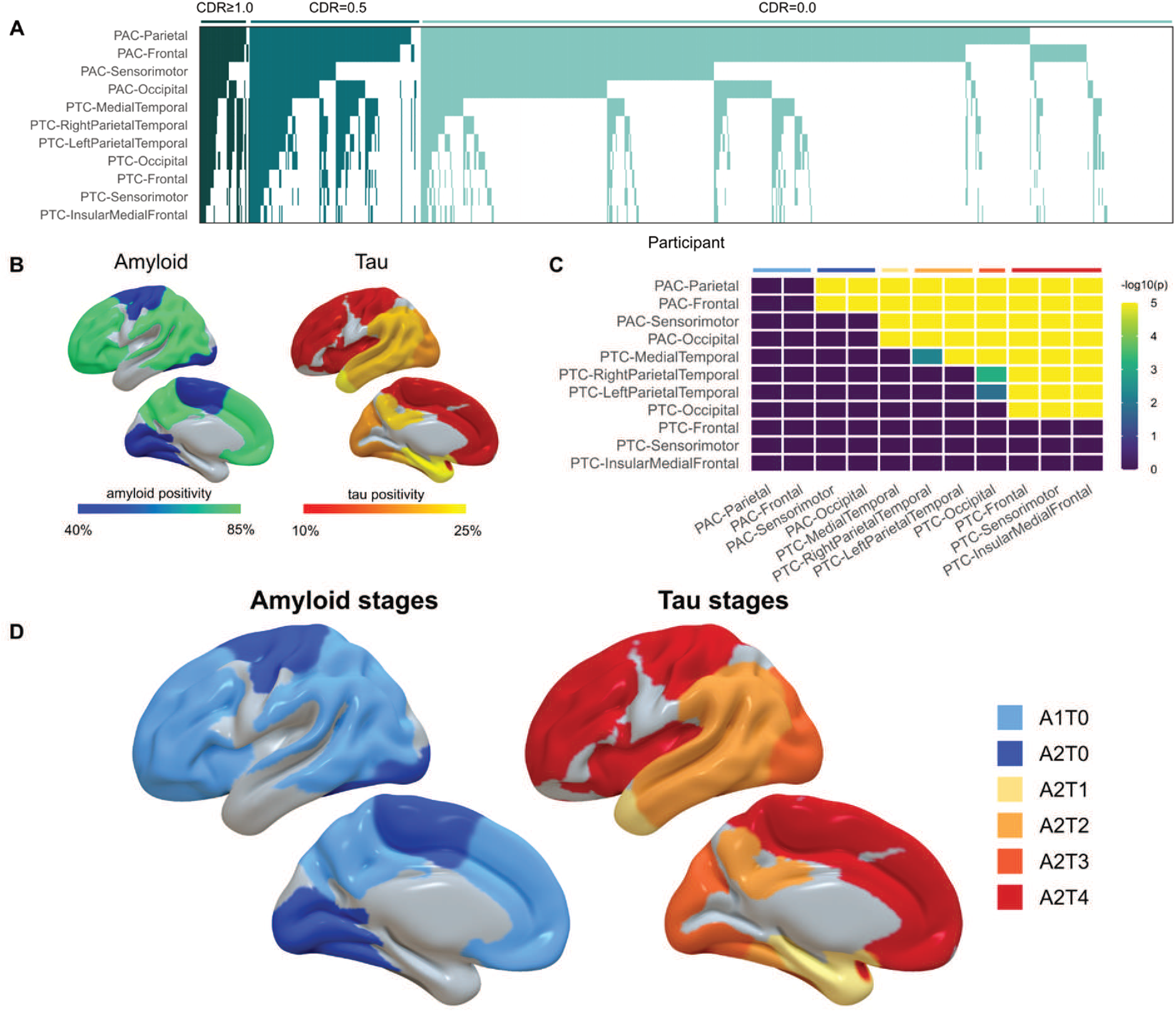
Development of the amyloid and tau staging model. **(A)** Ranking of PACs and PTCs by frequency of elevated pathology in training-ADS. Each column represents one participant, and filled cells represent suprathreshold W-scores. **(B)** Visualizations of the frequency of pathology across PACs (left) and PTCs (right) in training-ADS. **(C)** Results of the bootstrap algorithm for determining the number of stages. The cells are filled proportional to (the negative logarithm of) p-values of tests assessing whether cross-sectional frequencies of pathology are different across regions. **(D)** The proposed staging model showing the progression of amyloid (left) and tau (right).

We next applied a bootstrapping method to identify unique stages of AD pathology. This approach non-parametrically estimates which regions have significantly higher rates of positivity relative to others, informing the estimated steps in which pathology spreads across the brain. Unlike other similar approaches (*13, 17*), our method automatically determines the number of disease stages based on pairwise differences in the rate of pathology between regions. Application of our algorithm identified six stages of pathology, revealing a cascade beginning with diffuse amyloid pathology and followed by tau spread (Figure 3C & D). The first stage (A1T0) indicated broad amyloid uptake in canonical composite regions (PAC-Frontal, PAC-Parietal), while a second amyloid stage (A2T0) showed amyloid forming in sensory and motor areas (PAC-Occipital, PAC-Sensorimotor). The third stage marked the onset of tau pathology (A2T1) limited to medial temporal regions (PTC-MedialTemporal). The fourth stage (A2T2) reflected tau spread to bilateral temporal and parietal cortices (PTC-LeftParietalTemporal, PTC-RightParietalTamporal), while a fifth stage (A2T3) indicated further spread to the occipital lobe (PTC-Occipital). The final stage (A2T4) showed tau deposition in frontal, insular, and sensorimotor areas (PTC-Frontal, PTC-InsularMedialFrontal, PTC-Sensorimotor).

We replicated W-scoring and stage derivation in the validation dataset. The positivity-defined ordering of PACs and PTCs in the validation data was consistent with that of the training set, indicating a similar estimated temporal ordering of regions (Friedman test, χ^2^=22, df=2, p<0.001; Supplementary Figure 8A&B). The derived staging model also indicated a similar pathological cascade, though the number of stages and specific grouping of PACs and PTCs differed (Supplementary Figure 8C). Despite these differences, individual stage assignments were consistent across models (ARI=0.60; Supplementary Figure 8G). Results were similarly consistent when models were derived from fixed-tracer subsets of the validation data (Supplementary Figure 8D-G, ARI>0.57).

### Cross-dataset application and longitudinal stability of the staging model

We staged both the training-ADS and validation-ADS datasets using our model (Figure 4A, B). In the training-ADS cohort, most individuals (80.0%) had pathological distributions consistent with the staging model, while the remaining (20.0%) were marked as non-stageable (NS). The distribution of stageable and NS individuals was not associated with age, race, sex, or source dataset (all p>0.05). Of the stageable individuals, most exhibited amyloid pathology without tau (A1T0: 28.4%; A2T0: 29.0%), with a smaller proportion displaying tau pathology (A2T1: 2.3%; A2T2: 3.5%; A2T3: 1.9%, A2T4: 7.6%). A minority of individuals were labeled as negative for any pathology (A0T0: 7.3%). These A0T0 cases captured individuals with amyloid levels near the positivity threshold used by most source datasets (Figure 4C). Similar findings were observed in the validation-ADS cohort, where 84% of the individuals were stageable. The distribution of stages was comparable, with most individuals classified as A1T0 (18.3%) or A2T0 (38.6%), with fewer in tau stages (A2T1: 2.5%; A2T2: 2.4%; A2T3: 4.0%, A2T4: 14.6%), and a minority classified as no pathology (A0T0: 3.6%). Again, the distribution of stageable and NS presentations did not vary with age, sex, race, or dataset source (all p>0.05). Average SUVR maps stratified by stage showed PET uptakes consistent with the topography defined by the staging model (Figure 4E, F).

**Figure 4.**
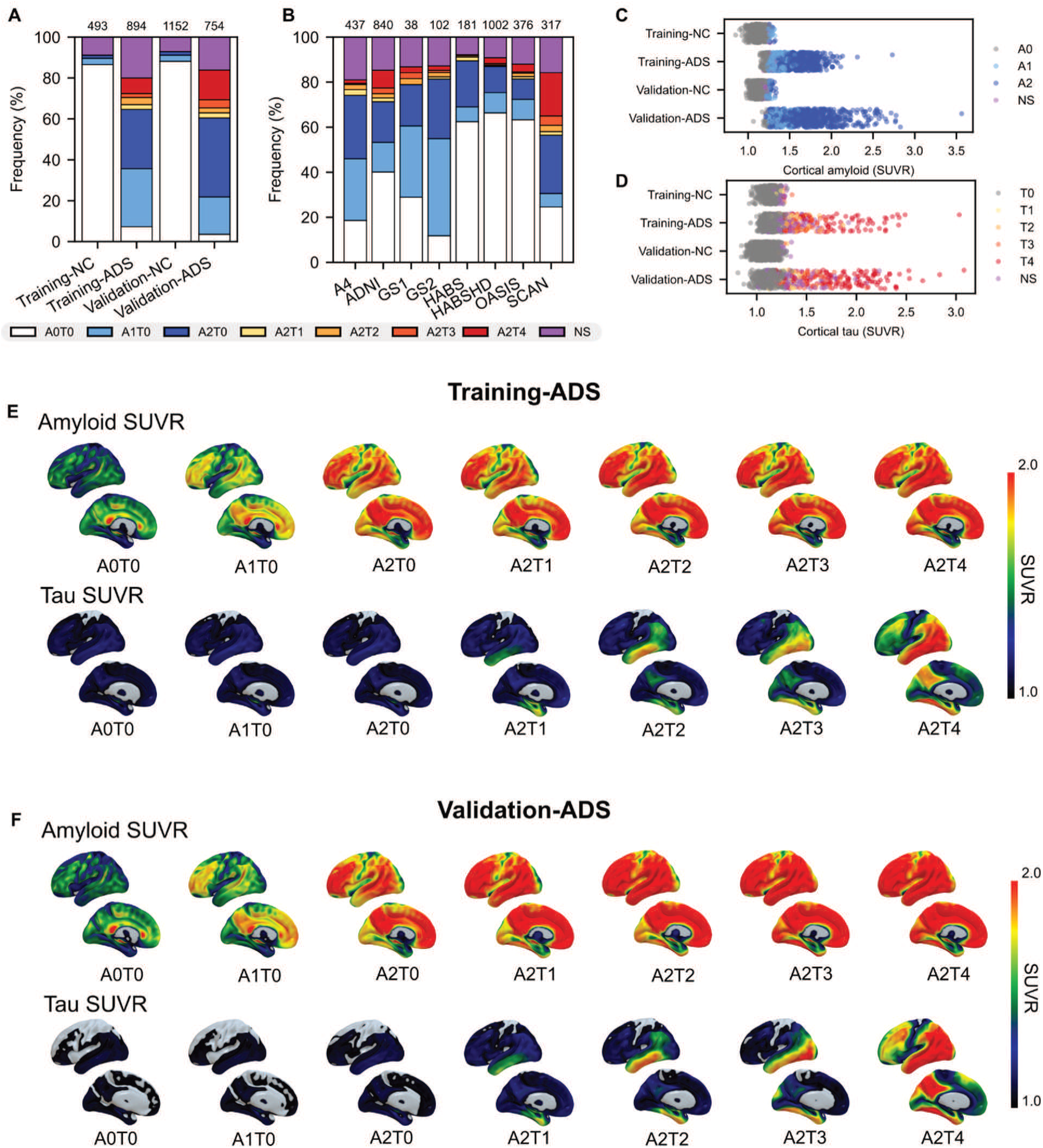
Application of the staging model. **(A)** Distribution of stages for all included participants stratified by training/validation split and disease status. **(B)** Distribution of stages for all included participants stratified for dataset of origin. **(C)** Scatter plot showing the distribution of cortical amyloid SUVR across study groups. **(D)** Scatter plot showing the distribution of cortical tau SUVR across study groups. For (C) and (D), observations are colored by amyloid and tau stage, respectively. **(E)** Images showing the average distribution of amyloid (top) and tau (bottom) SUVRs by staging group in training-ADS. **(F)** Same as (E), but for validation-ADS.

We used longitudinal imaging data to test whether staging assignments progressed in accordance with the inferred pathological cascade (Supplementary Figure 9). Of 406 training-ADS individuals with longitudinal data, most followed the staging model over time. A permutation test confirmed that stage transitions were longitudinally consistent with the model (p<0.001). Similar results were observed in 241 validation-ADS individuals with longitudinal imaging (permutation test: p<0.001).

### Staging captures dementia severity and predicts clinical progression

We next validated the relationship between staging and clinical and cognitive measures of AD progression. To establish a normative reference, NC individuals were staged alongside ADS participants (Figure 4A). Cross-sectionally, disease stages were associated with more severe cognitive impairment in the training data. Global Clinical Dementia Rating (CDR) status differed with disease stages (chi squared test, χ^2^=385.58, df=14, p<0.001; Figure 5A), with higher disease stages becoming more prevalent as dementia severity increased. Correspondingly, CDR Sum of Boxes (CDR-SB) scores worsened with increasing disease stage (one-way ANOVA, F(7, 1236)=50.98, p<0.001; Figure 5B, Supplementary Table 2). Impairment relative to A0T0 was detectable at A2T0 (Tukey test, p<0.01) but not at A1T0 (Tukey test, p=0.206). Similarly, Mini Mental State Examination MMSE scores worsened with increasing stages (one-way ANOVA, F(7, 1343)=174.97, p<0.001; Figure 5C, Supplementary Table 3) with significant decline first observable at A2T0 (Tukey test, p<0.001). Similar patterns were observed in validation data (Figure 5D-F, Supplementary Table 4, Supplementary Table 5). CDR-SB (one-way ANOVA, F(7, 1833)=206.3, p<0.001) and MMSE (one-way ANOVA, F(7, 1551)=38.5, p<0.001) scores worsened with disease stages, and CDR-SB deficits were first observable at A2T0 (Tukey test, p<0.001). However, statistically significant MMSE differences relative to A0T0 were not observed until A2T3 (p<0.001).

**Figure 5.**
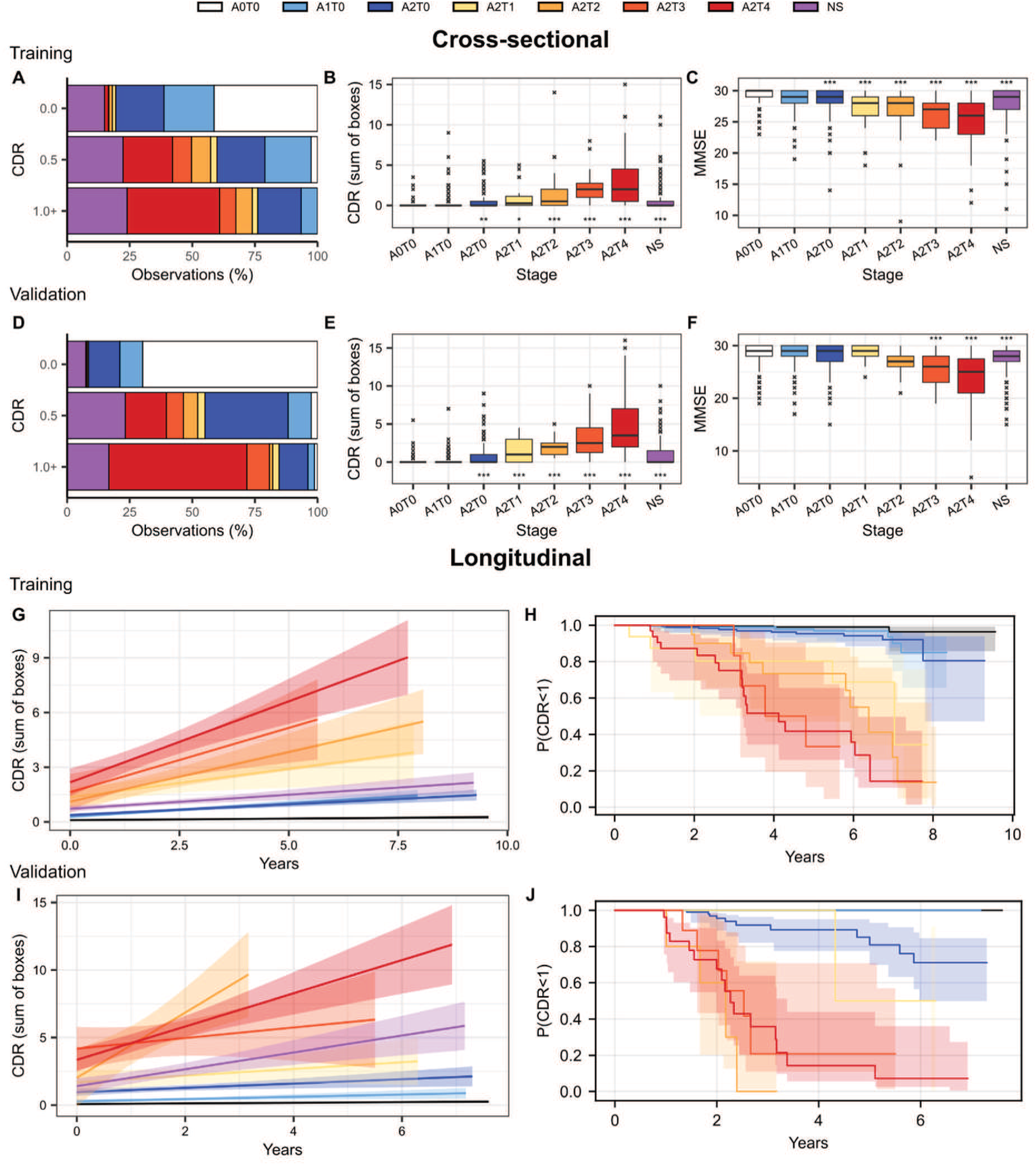
The association of disease stage with clinical and cognitive decline. **(A)** Distribution of disease stages against CDR status in the training set. **(B)** Boxplots comparing CDR-SB across disease stages (training). **(C**) Boxplot comparing MMSE scores across disease stages (training). **(D-F)** Same as (A-C) but for validation. **(G)** Fit lines from mixed effect models predicting longitudinal CDR-SB scores from baseline disease stage (training). **(H)** Kaplan-Meier survival curves showing the risk of dementia development for each disease stage (training). **(I-J)** Same as (G-H) but using the validation dataset. In (B), (C), (E), and (F), stars represent statistically significant differences from the A0T0 group (Tukey test after one-way ANOVA, *p<0.05, **p<0.01, ***p<0.001).

We also tested if stages predicted future cognitive decline. In the training dataset, baseline stages were associated with longitudinal CDR-SB trajectories (F=31.9, p<0.001; Figure 5G, Supplementary Table 6). Relative to A0T0, steeper decline was detectable for all stages (all p<0.01) except A1T0 (t-ratio=-2.695, df=1214.4, p=0.124). A1T0 & A2T0 had similar rates of decline (t-ratio=-0.846, df=1215.8, p=0.99), and all tau stages (A2T1-A2T4) declined more rapidly than A2T0 (all p<0.001). Significant differences between tau stages were only observed when comparing A2T4 to A2T1 (t-ratio=-4.1, df=1211.8, p<0.001) or A2T2 (t-ratio=-4.1, df=1218.6, p<0.001). MMSE decline also varied across stages (F=22.8, p<0.001; Supplementary Figure 10A, Supplementary Table 7). Relative to A0T0, faster decline was only detectable after A2T1 (t-ratio=6.9, df=1187.8, p<0.001). Furthermore, all tau stages showed similar rates of decline (all p>0.05), except for A2T4 declining faster than A2T2 (t-ratio=3.727, df=1174.8, p<0.01). These findings replicated in the validation dataset. CDR-SB trajectories differed by stage (F=45.5, p<0.001; Figure 5I, Supplementary Table 8), with significant differences first emerging at A2T0 (t-ratio=-5.225, df=619.4, p<0.001). All tau stages declined faster than amyloid stages (all p<0.05), and A2T1 showed slower decline than later tau stages (all p<0.01). MMSE trajectories similarly varied by stage (F=23.6, p<0.001; Supplementary Figure 10B, Supplementary Table 9), with accelerated decline first observable at A2T2 (t-ratio=4.5, df=495.7, p<0.001).

Lastly, we conducted survival analyses to test if baseline disease stage predicted conversion to mild or worse dementia (CDR≥1). In the training dataset, A2T0 individuals converted faster than A0T0 (log-rank statistic=7.0, p<0.001), while A1T0 individuals (log-rank statistic=0.058, p=0.058) did not. Individuals in tau stages progressed faster than individuals in amyloid stages (all p<0.001), though differences were not observed between tau stages (all p>0.05). The validation dataset showed similar patterns. A2T0 individuals progressed faster than A0T0 (log-rank statistic=28.82, p<0.001) and A1T0 individuals (log-rank statistic=5.88, p<0.05), whereas A1T0 did not differ from A0T0 (log-rank statistic=0.0, p=1.0).

### Non-stageable presentations reflect distinct tau-dominant patterns

NS presentations were subdivided into the following subgroups: individuals without amyloid but with tau (A0T+), individuals with first stage amyloid and tau (A1T+), and individuals with both amyloid stages and late-stage tau without medial temporal involvement (MTL-) (Supplementary Figure 11). All remaining NS cases reflected atypical late tau involvement (Atypical Tau). Across cohorts, most NS cases were MTL- presentations (training-ADS: 50.8%, validation-ADS: 56.2%, respectively), followed by Atypical Tau (training-ADS: 23.5%, validation-ADS: 31.4%). A1T+ cases were less frequent (training-ADS: 20.7%, validation-ADS: 12.4%), while A0T+ cases were rare (training-ADS: 5.0%, validation-ADS: 0%).

We ran additional models to compare stageable and NS individuals (Supplementary Table 12). In training data, we did not find age, sex, education, BMI, or *APOE* E4 carriership differences between these groups (all p>0.05). Cognitively, NS individuals showed an intermediate presentation with worse CDR-SB compared to the amyloid stages (NS vs. A1T0-A2T0: difference=0.493, p<0.05) but better than the tau stages (NS vs A2T1-A2T4: difference=- 1.439, p<0.001). The same was true for MMSE scores (NS vs. A1T0-A2T0: difference=-0.863, p<0.01; NS vs A2T1-A2T4: difference=1.829, p<0.001). Accounting for global amyloid and tau burden, we found that NS individuals had higher W-scores in PTC-MedialTemporal (difference=0.408, p<0.01), PTC-Sensorimotor (difference=0.454, p<0.05), and PTC-InsularMedialFrontal (difference=0.412, p<0.01) than the amyloid A1T0-A2T0 cases. No differences were found in factor uptakes between NS cases and A2T1-A2T4 cases. Similar results were observed in validation data (Supplementary Table 13).

### Comparison with Alzheimer’s Association biological and clinical stages

We contextualized our staging model relative to the Alzheimer’s Association biological and clinical staging frameworks (*1*), hereafter referred to as AA2024 Biological and AA2024 Clinical. Our proposed stages showed strong correspondence with AA-2024 Biological stages (Figure 6A). For each of our proposed stages, we found that most individuals were assigned to a single AA2024 Biological stage. Amyloid-only stages aligned with A+/T_2_- (A1T0: 94.5%; A2T0: 93.4%). Tau stages were more varied but mapped progressively to higher AA2024 Biological categories: A2T1 with A+/T_2MTL_+ (66.7%), A2T2 and A2T3 with A+/T_2MOD_+ (58.1% and 58.8%, respectively), and A2T4 with A+/T_2HIGH_+ (77.4%). We found a similar correspondence in the validation dataset (Figure 6D), although A2T3 cases were split approximately evenly between A+/T_2MOD_+ (40.0%) and A+/T_2HIGH_+ (40.0%).

**Figure 6.**
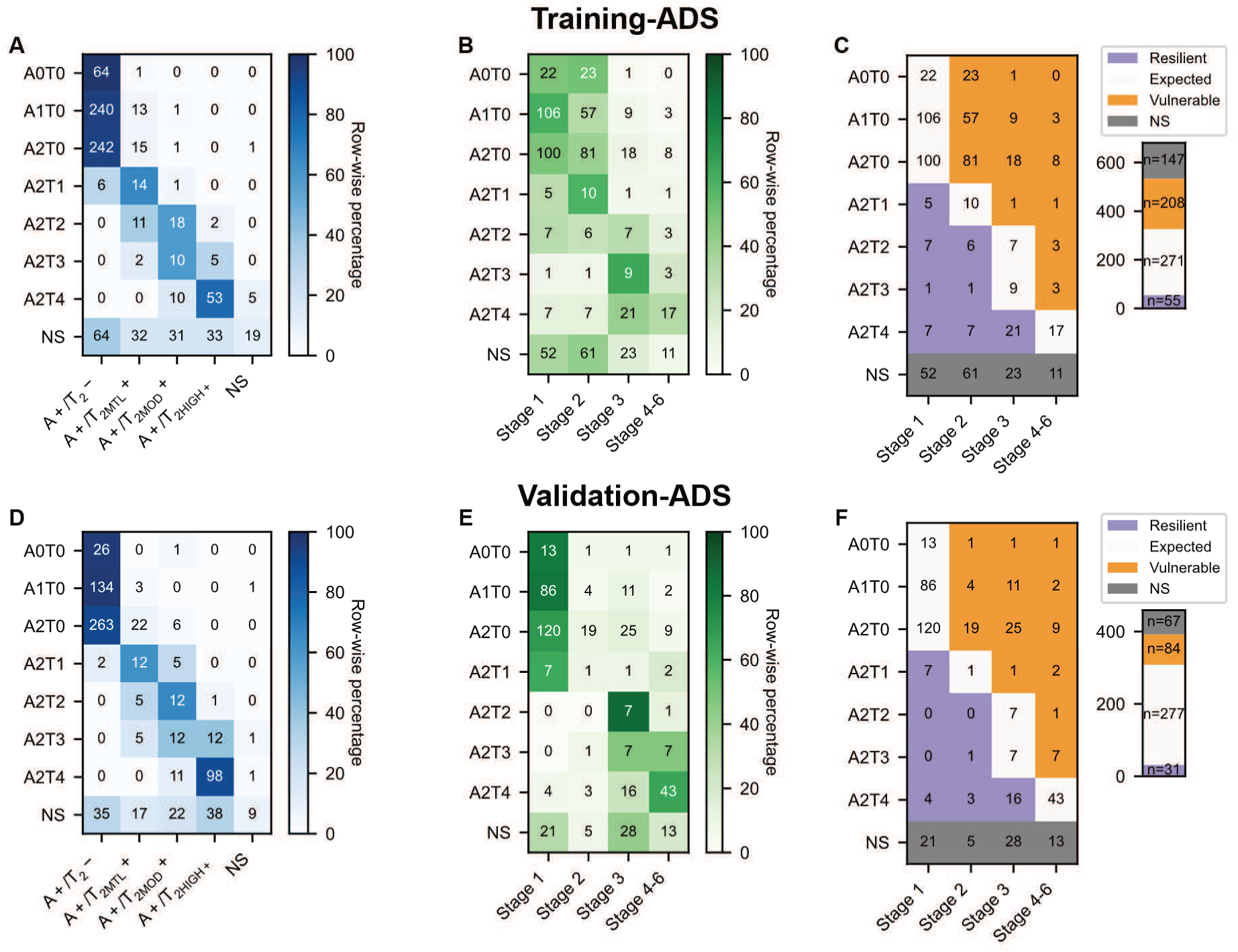
Comparison of the proposed staging model with frameworks proposed by the Alzheimer’s Association 2024 criteria. **(A)** Cross tabulation of proposed staging and AA-2024 Biological stages (training-ADS). **(B)** Cross tabulation of proposed staging and AA-2024 Clinical stages (training-ADS). **(C)** Heatmap showing the distribution of resilient (clinical stage lower than biological stage), vulnerable (clinical stage higher than biological stage), and expected (clinical stage matches biological stage) individuals (training-ADS) **(D-F)** Same as **(A-C)** but with validation-ADS.

For a subset of individuals with the required cognitive assessments (training-ADS: n=681, validation-ADS: n=459), we additionally assigned AA2024 Clinical stages. Alignment with AA2024 Clinical was less consistent (Figure 6B, E), indicating differential clinical susceptibility to AD pathology. Leveraging the AA2024 framework (*1*), we categorized individuals as expected, vulnerable (worse clinical stage than biologically predicted), and resilient (better clinical stage than biologically predicted; Figure 6C, Supplementary Table 14). In training-ADS, the plurality of individuals fit the expected trajectory (n=271, 39.8%), 30.5% (n=208) were classified vulnerable, and 8.0% (n=55) as resilient. The vulnerable group had lower tau burden than the expected group (difference=0.05 SUVR, p<0.05), with no differences in amyloid burden (difference=0.006, p=0.783) and gray matter volume (difference=0.006, p=0.999). As expected, MMSE scores were lower in the vulnerable group (difference=1.1, p<0.01). Vulnerable individuals were also older (difference=2.6 years, p<0.01), more frequently male (difference=12.6%, p<0.05), and had lower education (difference=0.623, p<0.05). In contrast, resilient individuals had lower gray matter volume (difference=0.012, p<0.001), higher amyloid burden (difference=0.193, p<0.001), higher tau burden (difference=0.350, p<0.001), and more frequent *APOE* E4 carriership (difference=17.1%, p<0.01) than expected individuals, with no differences in demographic variables. Replication in validation-ADS (Figure 6, Supplementary Table 15), confirmed that vulnerable individuals were older (difference=4.3 years, p<0.01) and had lower education (difference=1.1 years, p<0.001). No differences were found between the resilient and expected group in validation-ADS (all p>0.05).

## DISCUSSION

In this study, we developed a staging model for AD which captures the regional progression of amyloid and tau pathology. Using NMF, we identified four amyloid (PACs) and seven tau (PTCs) factors, encoding areas where pathology accumulates in a coordinated manner. Importantly, these factors were highly reproducible across independent datasets with varied PET tracers, scanners, and acquisition parameters, supporting their biological relevance. Leveraging these factors, we defined a six-stage model of disease progression. In our model, amyloid deposition begins in frontal and parietal cortex (A1T0) and extends to primary sensory and motor regions (A2T0). The formation of tau follows, originating in medial temporal regions (A2T1) and spreading externally to lateral temporoparietal (A2T2), occipital (A2T3), and widespread neocortical areas (A2T4). Disease stages coincided with clinically meaningful cognitive trajectories, both cross-sectionally and longitudinally. Crucially, our findings were generated using a bottom-up approach relying on data-driven methods rather than pre-specified frameworks. Our results provide an empirically informed model of disease progression which spans the entire development of pathological proteinopathies in AD. The model illuminates the biological progression which defines AD and can be applied for individual assessment of disease severity and prognostic risk.

Our results focused on regions of amyloid and tau accumulation extracted from high-dimensional image volumes with NMF. The PACs and PTCs were identified without *a priori* assumptions about which brain regions are susceptible to AD pathology and were not confined to conventional ROI boundaries. Instead, areas of interest were selected by empirically identifying the brain structures which tend to harbor pathology across a large sample. This approach contrasts with prior investigations which have focused on ROI analyses (*7, 12–15*), sometimes requiring manual selection of the regions that inform staging (*16–18*). As such, our results present a novel and complementary understanding of the development and spread of AD pathology.

PACs and PTCs define a low-dimensional representation of regional disease burden suitable for individualized assessments. PAC-Frontal and PAC-Parietal encompassed canonical regions typically used for automated amyloid quantification (*23*). The separation of this meta-region into multiple factors suggests heterogeneity in amyloid accumulation, consistent with prior work identifying clusters of individuals with frontal or parietal dominant pathology (*17, 25*). PAC-Occipital and PAC-Sensorimotor instead highlight regions previously shown to be final places of cortical amyloid deposition (*12*). Tau factorization resulted in seven factors, demonstrating greater spatial complexity. PTC-Medial Temporal corresponded to early Braak stages (I/II), PTC-LeftParietalTemporal and PTC-RightParietalTemporal highlighted areas of subsequent expansion (Braak III/IV), while PTC-Sensorimotor aligned with the final Braak stage

(VI). However, regions encompassed within Braak V were further partitioned into PTC-Occipital, PTC-Frontal, and PTC-InsularMedialFrontal, indicating refinement beyond the traditional Braak framework. Additionally, lateralized PTC-LeftParietalTemporal and PTC-RightParietalTemporal captured asymmetric tau deposition patterns, a known presentation often linked to language phenotypes and aggressive decline (*16, 26, 27*). These NMF-derived pathology patterns were observable across independent datasets and PET tracers, with differences primarily stemming from hierarchical merging of factors. This remarkable stability further supports the notion that PACs and PTCs capture biologically meaningful deposition patterns which can be leveraged for individualized quantification of AD pathology.

A central contribution of this work is the integration of amyloid and tau into a staging framework. Previous data-driven approaches have mainly focused on staging a single pathology (*7, 12–15*). AD is defined by the presence of amyloid and tau proteinopathies, and biological staging of the disease requires consideration of both pathologies (*1*). Importantly, we applied the same unsupervised methods for assessing amyloid and tau, not relying on previously defined ROIs, staging models, or cutoffs. Without enforcing any ordering, our results indicated that tau pathology occurs only after substantial amyloid involvement, consistent with the understanding that amyloidosis is nearly always a prerequisite for neocortical tauopathy (1). Regional rates of amyloid pathology exceeded those of tau, and isolated tau pathology without amyloid was rare.

Our methods indicated two stages of amyloid pathology: a first showing diffuse frontal, parietal (A1T0), and temporal pathology followed by a second with sensorimotor and occipital involvement (A2T0). This multistage amyloid progression contrasts with binary amyloid-positive classifications used in research (*1*) and clinic. Other staging works focused on amyloid progression have shown multistage amyloid progressions (*7, 12, 13*). Our A1T0 and A2T0 most closely resemble intermediate and late amyloid stages described previously (*12, 13*), though we did not observe distinct early cingulate-only or late striatal stages. In our work, cingulate cortices loaded onto both PAC-Frontal and PAC-Parietal, suggesting amyloid formation to be correlated with other broad cortical regions.

Despite the use of voxel-based analysis and incorporation of different datasets, our model was largely consistent with the four tau stages developed in our previous work (*15*). Our model also showed several key similarities with Braak staging, particularly the initial tau stage (A2T1) consisting of medial temporal pathology followed by expansion to broader parietal and temporal cortices (A2T2). Although, PTC-MedialTemporal included some regions external to entorhinal cortex and parts of the temporal pole, so it may be capturing regions tau accumulation in between Braak I/II and Braak III/IV. We further observed a separate occipital stage (A2T3) and a final stage for frontal, insular, and sensorimotor development (A2T4), patterns which diverge from the Braak model. This regional hierarchy of neocortical regions also differed from our previous work (*15*), which combined frontal and occipital tau into a single stage, and from other data-driven approaches to find tau stages (*14*). Variability in neocortical tau stages across the literature likely reflects cohort differences and biological heterogeneity, which may affect the inference of a single staging trajectory from cross-sectional data (*16, 26*).

Multiple analyses confirmed that cognition was associated with disease stage. Higher stages corresponded to worse baseline cognitive performance, accelerated cognitive decline and dementia progression. As expected, individuals with tau pathology (A2T1-A2T4) showed substantially greater impairment than those in amyloid-only stages (A1T0-A2T0). Interestingly, A2T0 individuals exhibited subtle cognitive decline relative to individuals without pathology (A0T0), whereas A1T0 did not. These results could indicate that advanced amyloid pathology may coincide with early cognitive vulnerability even prior to overt tau involvement. This is consistent with prior work showing differential cognitive trajectories associated with individuals in different amyloid stages (*7, 28*). Although statistical separation between A1T0 and A2T0 groups was limited, effect sizes for cognitive differences relative to controls were consistently higher for the A2T0 group, supporting potential utility for identification of fast and slow disease progressors.

Approximately 20% of individuals we assessed did not conform to the staging model (NS). This proportion is larger than previous staging studies of amyloid or tau, partly reflecting the additional granularity offered by our model. Modeling the spatial spread of both amyloid and tau allows for more potential types of NS presentations than models based on one pathology. Additionally, other staging work has shown evidence for sizable proportions of individuals with unique progressions that differ from the majority trend (*16*). Further investigation of the NS cases showed evidence of heterogeneous deposition patterns. Of the NS individuals, around 80% were due to atypical tau patterns, particularly medial-temporal sparing presentations consistent with known AD subtypes (*29*). A small number of individuals showed tau pathology with no amyloid or atypical amyloid presentations. NS showed a moderate cognitive phenotype, with cross-sectional and longitudinal impairment generally higher than those with only amyloid pathology but less than those that reached the tau stage.

Comparison with AA2024 biological and clinical stages (*1*) demonstrated concordance with biological stages, with few individuals displaying a change of more than one stage label. However, clinical stages were more variable. Consistent with previous work, a sizable proportion of individuals appeared clinically vulnerable or resilient relative to their biological stage (*30, 31*). We specifically identified a higher proportion of vulnerable individuals, who were characterized by older age and lower educational attainment, replicating some of the associations reported in prior work (*31*).

Several limitations should be noted. First, stage ordering was inferred from cross-sectional patterns of regional uptake rather than directly observed longitudinal trajectories. Although cross-sectional disease modeling under monotonic accumulation assumptions is well-established, it mixes information from individuals who may have unique spatial progressions. Longitudinal data would provide a more direct assessment of within-individual change. However, large-scale datasets with sufficiently long follow-up to detect measurable amyloid and tau accumulation are currently limited. Second, we relied on binarization of regions to define individualized measures of regional pathology, which introduces threshold sensitivity and may contribute to classification variability, particularly for individuals near positivity cutoffs. Third, the study population was majority white and well educated despite including in the validation dataset the HABS-HD cohort, which has a larger proportion of black and Hispanic individuals. However, we found no evidence that the rate of conformity to the staging model was associated with race. Finally, we mixed data from multiple sources in our analysis. While this design enhanced stability and external validity, differences in acquisition, reconstruction and initial preprocessing may introduce residual nuisance variability.

In summary, unsupervised voxel-wise analyses identified reproducible spatial patterns of coordinated amyloid and tau deposition that inform a unified six-stage biological framework for AD. This model operationalizes hierarchical proteinopathy involvement using in vivo PET imaging, supports individualized assessment of disease burden, and provides a data-driven foundation for biological stratification and prognostic evaluation. As disease-modifying therapies continue to evolve, biologically grounded staging frameworks may play an increasingly important role in precision characterization of Alzheimer’s disease.

## MATERIALS AND METHODS

### Study Design

The aim of this study was to derive a data-driven model which stages both amyloid and tau pathology suitable for application in AD. We pursued this aim by using a retrospective, observational design of individuals with AD and healthy controls. We aimed to maximize the sample size by sourcing data from large, publicly available, multisite imaging studies of AD. No explicit sample size calculation was applied. We considered for inclusion all individuals who received amyloid- and tau-PET imaging (within 1 year). A T1-weighted MRI scan was also required for image processing purposes. We separated included individuals into a training set (for model development and application) and validation set (for replication of findings). The training set consisted of people scanned with FBP and FTP, while the validation set was composed of three subsets which were unique in their amyloid/tau tracer pairs utilized (validation-A: FBB/FTP, validation-B: FBB/PI2620, and validation-C: PIB/FTP). Individuals were additionally assigned to one of two disease status groups: ADS (positive for amyloid pathology) or NC (negative for amyloid pathology, negative for tau pathology, and no dementia), with dementia status assessed by the Clinical Dementia Rating ® (CDR) (*32*). The assignment of individuals into disease status and training/validation groups resulted in four primary sub-cohorts referred to through the study: training-ADS, training-NC, validation-ADS, and validation-NC. Exclusion criteria for the study were (a) not having the required imaging data, (b) not having PET tracers matching the training or validation sets, (c) being neither ADS nor NC, or (d) failing quality control checks after image preprocessing.

### Data Sources

We included data from 8 different sources: Anti-Amyloid Treatment in Asymptomatic Alzheimer’s (A4) (*33*), Alzheimer’s Disease Neuroimaging Initiative (ADNI), Alzheimer’s Prevention Initiative Generation Program Studies 1 (GS1) and 2 (GS2) (*34*), Harvard Aging Brain Study (HABS) (*35*), Health and Aging Brain Study: Health Disparities (HABS-HD) (*36*), Open Access Series of Imaging Studies (OASIS) (*37*), and the National Alzheimer’s Coordinating Center Standardized Centralized Alzheimer’s & Related Dementias Neuroimaging (SCAN). HABS data came from Public Data Release v2.0, accessed in September 2024 via http://habs.mgh.harvard.edu/. For HABS-HD, we accessed Release 5. All participants provided informed written consent for participating.

Some data used in the preparation of this article were obtained from the ADNI database (adni.loni.usc.edu). The ADNI was launched in 2003 as a public-private partnership, led by Principal Investigator Michael W. Weiner, MD. The primary goal of ADNI has been to test whether serial magnetic resonance imaging (MRI), PET, other biological markers, and clinical and neuropsychological assessment can be combined to measure the progression of mild cognitive impairment (MCI) and early AD.

### Image Acquisition & Processing

Details on PET image acquisition from each dataset are provided in the Supplementary Material. All PET images (amyloid and tau) were preprocessed with a volumetric pipeline to generate SUVR images in a common space (see Supplementary Material). We processed as many sessions (unique imaging visits with amyloid- and tau-PET imaging) as available for all subjects. Measures of global amyloid and tau burden were derived from volume-weighted averages of meta-ROIs using regions suggested by the ADNI Imaging Core (listed in Supplementary Table 1). We followed previous approaches for identifying the MUSE ROIs which corresponded to FreeSurfer ones (*38*).

### Biomarkers

We assigned amyloid- and tau-positivity to all individuals to determine the presence of AD (*1*). If available (91.2% of included individuals), we selected the assessment of amyloid-positivity provided by the dataset (see Supplementary Material). For those with missing values, we imputed amyloid-positivity via a receiver operating characteristic (ROC) analysis. For each amyloid tracer (across studies), we identified the cutoff in our computed global amyloid composite which provided the best accuracy for classifying the amyloid-positivity of those without missing data (cutoffs: FBP=1.22, FBB=1.26, PIB=1.25). These cutoffs were used to assign amyloid-positivity in those subjects with missing data. To ensure the NC group was free of AD-pathology, a small number of individuals with high global amyloid SUVR (>1.35, n=13) were manually marked as amyloid-positive. As tau-positivity was generally not provided by the source datasets, we derived our own assessment for all subjects. For each tau tracer, we fit a 2-component Gaussian Mixture Model to the distribution of global tau SUVRs. We identified the positivity threshold as the intersection of the two components (cutoffs: FTP=1.36, P26=1.27).

### Cognitive Assessments

The Global Score of the CDR (*32*) assessment was used to determine dementia status (0=no dementia, 0.5=very mild dementia, 1=mild dementia, 2=moderate dementia, 3=severe dementia). The Sum of Box score (CDR-SB) was used as a finer grained measure of clinical impairment. The Mini-Mental State Exam (MMSE) total score was also used as a measure of global cognitive impairment (*39*). For purposes of clinical staging (described below), we computed a modified version of the Preclinical Alzheimer Cognitive Composite (PACC) (*40*) for subjects with available neuropsychological assessments (see Supplementary Material). For cross-sectional analyses, clinical assessments were only included if they were taken within 1 year of the mean date of acquisitions for amyloid- and tau-PET. Longitudinal analyses included all clinical assessments following the baseline imaging session.

### Non-negative Matrix Factorization (NMF)

A detailed description of NMF formulation is provided in Supplementary Methods. We constructed input data matrices for NMF consisting of vectorized amyloid- and tau-PET images in 1mm MNI space. Our primary NMF model was trained using imaging data from the training set. NMF was applied to amyloid and tau in separate models to identify factors which capture population-wide covariance for each pathology. A model selection procedure was applied to select the rank of NMF dimensionality reduction (i.e., number of factors) for amyloid and tau (independently). Following the rank selection, factors not corresponding to cortical gray matter regions were manually filtered out. For subsequent analyses, we calculated the SUVR uptake within each NMF factor for all individuals across cohorts.

In replication experiments, we applied NMF to the validation data using the rank parameters selected from the training NMF data. We also tested NMF on the subsets of the validation set (validation-A, validation-B, validation-C) to check for model stability across tracers. Solution similarity was evaluated by comparing the inner product of matched components evaluated in the validation set with that of the training set.

### Staging development

We derived a staging model for amyloid and tau informed by the cross-sectional distribution of pathology across PACs and PTCs in the training cohort. We first converted SUVRs in PACs and PTCs into W-scores, capturing their normative deviation from a control population (training-NC) (*15, 24*). Further details on W-score computation are provided in the Supplementary Material. W-scores within PACs and PTCs were thresholded at 2.5 to create regional binary assessments of amyloid- and tau-pathology. The binarized W-score data were passed to bootstrapping algorithm for grouping regional pathology-positivity distributions into a progressive staging model (*15*). Further details about this method are described in the Supplementary Material. Following the assignment of PACs and PTCs to stages, we staged all individuals (training and validation) by applying the estimated staging model. Individuals were labeled as positive for a given stage if they were positive for pathology in one of the regions contributing to that stage. Additionally, staging was cumulative such that individuals had to be positive for all prior stages to meet criteria for a later stage. Individuals with pathology distributions not conforming to the staging model were labeled as NS (non-stageable).

To investigate the reproducibility of our approach to staging, we repeated the estimation of our staging model in the validation cohort. W-scores in the validation set were calculated separately for each subset (A, B, and C) to ensure the models were tracer-specific. The staging model was refit using the whole validation dataset and its subsets individually. We compared the similarity of stage assignment under different staging schemes using the Adjusted Rand Index (ARI), a measure of clustering similarity taking on values between -1 and 1 (1 indicating perfect alignment, 0 indicating random similarity, and negative values indicating worse-than-random performance).

### Alzheimer’s Association biological and clinical staging

Our data-driven approach to staging amyloid and tau was compared to consensus recommendations for biological and clinical staging proposed by the Alzheimer’s Association (AA) and National Institute on Aging (NIA) (*1*), which we refer to as AA2024. The AA2024 Biological system proposes four hierarchical stages encompassing elevated amyloid (A+/T_2_-), elevated medial temporal tau (A+/T_2MTL_+), moderate neocortical tau (A+/T_2MOD_+), and high neocortical tau (A+/T_2HIGH_+). In our analysis, the first stage was equivalent to the amyloid-positivity assessment used in our analyses for determining ADS inclusion. For the tau stages, we followed a previous approach for defining medial temporal and neocortical regions of interest (*30*), translating their FreeSurfer regions of interest into MUSE alternatives (*38*). Summary SUVRs for these regions were calculated by taking volume-weighted regional averages. To match the approach to regional positivity used for PACs and PTCs, we calculated W-scores for the medial temporal and neocortical regions. T_2MTL_+ and T_2MOD_+ were assessed by having a W-score exceeding 2.5 in the respective ROI. The cutoff for T_2HIGH_+ was set at the median value of scores for those who were T_2MOD_+ (*30*). AA2024 biological stage labels were assigned hierarchically such that individuals at a given stage had to meet criteria for all prior stages, with non-conforming individuals labeled as NS.

We further classified into individuals into AA2024 Clinical, the staging model for clinical impairment (*1*). Individuals who were CDR=0 were labeled as Stage 1 (asymptomatic) or Stage 2 (transitional decline). Stage 2 individuals were distinguished from Stage 1 individuals using a modified approach to determine subtle cognitive impairment (SCI) (*30, 41*). Briefly, we identified individuals who had remained CDR=0 for at least 5 years after baseline and identified the 10^th^ percentile of PACC scores as the SCI cutoff. Individuals were labeled as Stage 3 (cognitive impairment with early functional impact) if they were CDR=0.5 and below the SCI cutoff, while Stage 4-6 (dementia with mild, moderate, or severe functional impact) was identified by CDR ≥1.0.

We used our proposed staging model and the AA2024 Clinical stages to identify individuals who had more or less advanced clinical profiles relative to their biological presentation (*30, 31*). AA2024 defines an expected progression for clinical and biological staging (Stage 1: A+/T_2_-; Stage 2: A+/T_2MTL_+; Stage 3: A+/T_2MOD_+, Stage 4-6: A+/T_2HIGH_+). Our proposed amyloid and tau stages were matched to AA2024 Biological stages (based on cross-tabulations of the two staging approaches) and used to inform the expected AA2024 Clinical stage. Individuals with non-missing AA2024 Clinical stages were labeled as either expected (expected clinical stage given biological stage), resilient (less advanced clinical stage than biological stage), vulnerable (more advanced clinical stage than biological stage), or NS (biological stage not-conforming to the staging model).

### Statistical Analysis

All analyses were conducted with α=0.05. The training and validation cohorts were evaluated for differences using t-tests for continuous variables and Chi-squared tests for categorical variables. The ordering of PACs and PTCs (ranked by frequency of pathology) in training and validation sets was compared using a Friedman test. The longitudinal stability of stage assignment was evaluated with a permutation test (see Supplementary Material).

One-way ANOVAs were used to compare CDR-SB and MMSE scores across stages (with Tukey post-hoc testing). Longitudinal cognitive score trajectories were modeled using linear mixed-effects models (fixed effects: age, sex, stage, time, stage×time; random slopes per subject) and compared across stages using estimated marginal means testing with FDR correction (*42*). Kaplan-Meier survival analyses were used to model risk of dementia development (CDR ≥1.0) for each stage, and post-hoc, FDR-corrected log-rank tests were used to compare survival curves against stages.

One-way ANCOVAs (adjusted for age and sex, with post-hoc Tukey testing) and chi-squared tests were used to compare stage conformity (stageable vs. NS) and cognitive susceptibility (expected vs. resilient vs. vulnerable) groups. When comparing stageable and NS individuals, regional pathology (the SUVR within each PAC and PTC) was compared after regressing out global amyloid or tau burden to adjust for the global intensity of pathology.

## Data Availability

All data were accessed and downloaded between August 2024 and January 2025. All data sources used for this study (A4, ADNI, GS1, GS2, HABS, HABS-HD, OASIS, NACC-SCAN) are publicly available datasets which were accessible following formal applications and data usage agreements. All code written for this manuscript will be shared online (https://github.com/sotiraslab/earnest_nmf_ad_staging).

## List of Supplementary Materials

Supplementary Methods

Fig S1-S11

Tables S1-S15

References (43–60)

## Acknowledgments

Research reported in this publication was supported by National Institute on Aging of the National Institutes of Health under award number R01AG067103. The content is solely the responsibility of the authors and does not necessarily represent the official views of the National Institutes of Health.

Computations were performed using the facilities of the Washington University Research Computing and Informatics Facility, which were partially funded by NIH grants S10OD025200, 1S10RR022984-01A1 and 1S10OD018091-01. Computations were performed using the facilities of the Washington University Research Computing and Informatics Facility (RCIF). The RCIF has received funding from NIH S10 program grants: 1S10OD025200-01A1 and 1S10OD030477-01.

The A4 Study was a secondary prevention trial in preclinical Alzheimer’s disease, aiming to slow cognitive decline associated with brain amyloid accumulation in clinically normal older individuals. The A4 Study was funded by a public-private-philanthropic partnership, including funding from the National Institutes of Health-National Institute on Aging, Eli Lilly and Company, Alzheimer’s Association, Accelerating Medicines Partnership, GHR Foundation, an anonymous foundation, and additional private donors, with in-kind support from Avid Radiopharmaceuticals, Cogstate, Albert Einstein College of Medicine and the Foundation for Neurologic Diseases.The companion observational Longitudinal Evaluation of Amyloid Risk and Neurodegeneration (LEARN) Study was funded by the Alzheimer’s Association and GHR Foundation. The A4 and LEARN Studies were led by Dr. Reisa Sperling at Brigham and Women’s Hospital, Harvard Medical School, and Dr. Paul Aisen at the Alzheimer’s Therapeutic Research Institute (ATRI) at the University of Southern California. The A4 and LEARN Studies were coordinated by ATRI at the University of Southern California, and the data are made available under the auspices of Alzheimer’s Clinical Trial Consortium through the Global Research & Imaging Platform (GRIP). The complete A4 Study Team list is available on: https://www.actcinfo.org/a4-study-team-lists/. We would like to acknowledge the dedication of the study participants and their study partners who made the A4 and LEARN Studies possible.

Data collection and sharing for the Alzheimer’s Disease Neuroimaging Initiative (ADNI) is funded by the National Institute on Aging (National Institutes of Health Grant U19AG024904). The grantee organization is the Northern California Institute for Research and Education. In the past, ADNI has also received funding from the National Institute ofBiomedical Imaging and Bioengineering, the Canadian Institutes of Health Research, andprivate sector contributions through the Foundation for the National Institutes of Health(FNIH) including generous contributions from the following: AbbVie, Alzheimer’s Association;Alzheimer’s Drug Discovery Foundation; Araclon Biotech; BioClinica, Inc.; Biogen; Bristol-Myers Squibb Company; CereSpir, Inc.; Cogstate; Eisai Inc.; Elan Pharmaceuticals, Inc.; EliLilly and Company; EuroImmun; F. Hoffmann-La Roche Ltd and its affiliated companyGenentech, Inc.; Fujirebio; GE Healthcare; IXICO Ltd.; Janssen Alzheimer ImmunotherapyResearch & Development, LLC.; Johnson & Johnson Pharmaceutical Research &Development LLC.; Lumosity; Lundbeck; Merck & Co., Inc.; Meso Scale Diagnostics, LLC.;NeuroRx Research; Neurotrack Technologies; Novartis Pharmaceuticals Corporation; PfizerInc.; Piramal Imaging; Servier; Takeda Pharmaceutical Company; and Transition Therapeutics.

The Generation Program was supported by Novartis Pharma AG, Basel, Switzerland and Amgen, Thousand Oaks, CA, USA, in collaboration with the Banner Alzheimer’s Institute located in Phoenix, AZ, USA. Generation Study 1 was supported by funding from the National Institute on Aging (UF1 AG046150), part of the National Institutes of Health, as well as the Alzheimer’s Association, FBRI, GHR Foundation and Banner Alzheimer’s Foundation. We thank the staff and investigators of the studies as well as the participants and their study partners, whose help and participation made this work possible.

Data used in the preparation of this article were obtained from the Harvard Aging Brain Study (HABS P01AG036694 https://habs.mgh.harvard.edu/). The HABS study was launched in 2010, funded by the National Institute on Aging. and is led by principal investigators Reisa A. Sperling MD and Keith A. Johnson MD at Massachusetts General Hospital/Harvard Medical School in Boston, MA.

Research reported in this publication as part of the Health and Aging Brain Study: Health Disparities (HABS-HD) was supported by the National Institute on Aging of the National Institutes of Health under Award Numbers R01AG054073, R01AG058533, R01AG070862, P41EB015922 and U19AG078109. The content is solely the responsibility of the authors and does not necessarily represent the official views of the National Institutes of Health.

Data were provided (in part) by OASIS (OASIS-3: Longitudinal Multimodal Neuroimaging: Principal Investigators: T. Benzinger, D. Marcus, J. Morris; NIH P30 AG066444, P50 AG00561, P30 NS09857781, P01 AG026276, P01 AG003991, R01 AG043434, UL1 TR000448, R01 EB009352. AV-45 doses were provided by Avid Radiopharmaceuticals, a wholly owned subsidiary of Eli Lilly.)

The NACC database is funded by NIA/NIH Grant U24 AG072122. NACC data are contributed by the NIA-funded ADRCs: P30 AG062429 (PI James Brewer, MD, PhD), P30 AG066468 (PI Oscar Lopez, MD), P30 AG062421 (PI Teresa Gomez-Isla, MD), P30 AG066509 (PI Thomas Grabowski, MD), P30 AG066514 (PI Mary Sano, PhD), P30 AG066530 (PI Helena Chui, MD, Arthur Toga, PhD), P30 AG066507 (PI Marilyn Albert, PhD), P30 AG066444 (PI David Holtzman, MD), P30 AG066518 (PIs Lisa Silbert, MD, Miranda Lim, MD, PhD), P30 AG066512 (PI Thomas Wisniewski, MD), P30 AG066462 (PI Scott Small, MD), P30 AG072979 (PI David Wolk, MD), P30 AG072972 (PIs Charles DeCarli, MD, Rachel Whitmer, PhD), P30 AG072976 (PI Andrew Saykin, PsyD), P30 AG072975 (PI Julie Schneider, MD, MS), P30 AG072978 (PI Ann McKee, MD), P30 AG072977 (PI Robert Vassar, PhD), P30 AG066519 (PI Joshua Grill, PhD), P30 AG062677 (PIs Brad Boeve, MD, Ronald Petersen, MD, PhD), P30 AG079280 (PI Jessica Langbaum, PhD), P30 AG062422 (PI Gil Rabinovici, MD), P30 AG066511 (PI Allan Levey, MD, PhD), P30 AG072946 (PI Linda Van Eldik, PhD), P30 AG062715 (PI Sanjay Asthana, MD, FRCP), P30 AG072973 (PI Russell Swerdlow, MD), P30 AG066506 (PIs Glenn Smith, PhD, ABPP, David Lowenstein, PhD, Ranjan Duara, MD), P30 AG066508 (PIs Stephen Strittmatter, MD, PhD, Christopher Van Dyck, MD), P30 AG066515 (PI Victor Henderson, MD, MS), P30 AG072947 (PI Suzanne Craft, PhD), P30 AG072931 (PI Henry Paulson, MD, PhD), P30 AG066546 (PIs Sudha Seshadri, MD, Gladys Maestre, MD, PhD), P30 AG086401 (PI Erik Roberson, MD, PhD), P30 AG086404 (PI Gary Rosenberg, MD), P30 AG086403 (PI Angela Jefferson, PhD), P30 AG072958 (PIs Heather Whitson, MD, Gwenn Garden, MD, PhD), P30 AG072959 (PI Jagan Pillai, MD, PhD), P30 AG092752 (Ihab Hajjar, MD, MS).

SCAN is a multi-institutional project that was funded as a U24 grant (AG067418) by the National Institute on Aging in May 2020. Data collected by SCAN and shared by NACC are contributed by the NIA-funded ADRCs as follows: Arizona Alzheimer’s Center - P30 AG072980 (PI: Eric Reiman, MD); R01 AG069453 (PI: Eric Reiman (contact), MD); P30 AG019610 (PI: Eric Reiman, MD); and the State of Arizona which provided additional funding supporting our center; Boston University - P30 AG013846 (PI Neil Kowall MD); Cleveland ADRC - P30 AG062428 (James Leverenz, MD); Cleveland Clinic, Las Vegas – P20AG068053; Columbia - P50 AG008702 (PI Scott Small MD); Duke/UNC ADRC – P30 AG072958; Emory University - P30AG066511 (PI Levey Allan, MD, PhD); Indiana University - R01 AG19771 (PI Andrew Saykin, PsyD); P30 AG10133 (PI Andrew Saykin, PsyD); P30 AG072976 (PI Andrew Saykin, PsyD); R01 AG061788 (PI Shannon Risacher, PhD); R01 AG053993 (PI Yu-Chien Wu, MD, PhD); U01 AG057195 (PI Liana Apostolova, MD); U19 AG063911 (PI Bradley Boeve, MD); and the Indiana University Department of Radiology and Imaging Sciences; Johns Hopkins - P30 AG066507 (PI Marilyn Albert, Phd.); Mayo Clinic - P50 AG016574 (PI Ronald Petersen MD PhD); Mount Sinai - P30 AG066514 (PI Mary Sano, PhD); R01 AG054110 (PI Trey Hedden, PhD); R01 AG053509 (PI Trey Hedden, PhD); New York University - P30AG066512-01S2 (PI Thomas Wisniewski, MD); R01AG056031 (PI Ricardo Osorio, MD); R01AG056531 (PIs Ricardo Osorio, MD; Girardin Jean-Louis, PhD); Northwestern University - P30 AG013854 (PI Robert Vassar PhD); R01 AG045571 (PI Emily Rogalski, PhD); R56 AG045571, (PI Emily Rogalski, PhD); R01 AG067781, (PI Emily Rogalski, PhD); U19 AG073153, (PI Emily Rogalski, PhD); R01 DC008552, (M.-Marsel Mesulam, MD); R01 AG077444, (PIs M.-Marsel Mesulam, MD, Emily Rogalski, PhD); R01 NS075075 (PI Emily Rogalski, PhD); R01 AG056258 (PI Emily Rogalski, PhD); Oregon Health and Science University - P30 AG008017 (PI Jeffrey Kaye MD); R56 AG074321 (PI Jeffrey Kaye, MD); Rush University - P30 AG010161 (PI David Bennett MD); Stanford – P30AG066515; P50 AG047366 (PI Victor Henderson MD MS); University of Alabama, Birmingham – P20; University of California, Davis - P30 AG10129 (PI Charles DeCarli, MD); P30 AG072972 (PI Charles DeCarli, MD); University of California, Irvine - P50 AG016573 (PI Frank LaFerla PhD); University of California, San Diego - P30AG062429 (PI James Brewer, MD, PhD); University of California, San Francisco - P30 AG062422 (Rabinovici, Gil D., MD); University of Kansas - P30 AG035982 (Russell Swerdlow, MD); University of Kentucky - P30 AG028283-15S1 (PIs Linda Van Eldik, PhD and Brian Gold, PhD); University of Michigan ADRC - P30AG053760 (PI Henry Paulson, MD, PhD) P30AG072931 (PI Henry Paulson, MD, PhD) Cure Alzheimer’s Fund 200775 - (PI Henry Paulson, MD, PhD) U19 NS120384 (PI Charles DeCarli, MD, University of Michigan Site PI Henry Paulson, MD, PhD) R01 AG068338 (MPI Bruno Giordani, PhD, Carol Persad, PhD, Yi Murphey, PhD) S10OD026738-01 (PI Douglas Noll, PhD) R01 AG058724 (PI Benjamin Hampstead, PhD) R35 AG072262 (PI Benjamin Hampstead, PhD) W81XWH2110743 (PI Benjamin Hampstead, PhD) R01 AG073235 (PI Nancy Chiaravalloti, University of Michigan Site PI Benjamin Hampstead, PhD) 1I01RX001534 (PI Benjamin Hampstead, PhD) IRX001381 (PI Benjamin Hampstead, PhD); University of New Mexico - P20 AG068077 (Gary Rosenberg, MD); University of Pennsylvania - State of PA project 2019NF4100087335 (PI David Wolk, MD); Rooney Family Research Fund (PI David Wolk, MD); R01 AG055005 (PI David Wolk, MD); University of Pittsburgh - P50 AG005133 (PI Oscar Lopez MD); University of Southern California - P50 AG005142 (PI Helena Chui MD); University of Washington - P50 AG005136 (PI Thomas Grabowski MD); University of Wisconsin - P50 AG033514 (PI Sanjay Asthana MD FRCP); Vanderbilt University – P20 AG068082; Wake Forest - P30AG072947 (PI Suzanne Craft, PhD); Washington University, St. Louis - P01 AG03991 (PI John Morris MD); P01 AG026276 (PI John Morris MD); P20 MH071616 (PI Dan Marcus); P30 AG066444 (PI John Morris MD); P30 NS098577 (PI Dan Marcus); R01 AG021910 (PI Randy Buckner); R01 AG043434 (PI Catherine Roe); R01 EB009352 (PI Dan Marcus); UL1 TR000448 (PI Brad Evanoff); U24 RR021382 (PI Bruce Rosen); Avid Radiopharmaceuticals / Eli Lilly; Yale - P50 AG047270 (PI Stephen Strittmatter MD PhD); R01AG052560 (MPI: Christopher van Dyck, MD; Richard Carson, PhD); R01AG062276 (PI: Christopher van Dyck, MD); 1Florida - P30AG066506-03 (PI Glenn Smith, PhD); P50 AG047266 (PI Todd Golde MD PhD)

## Author contributions

Conceptualization: TWE, AS

Data curation: TWE, AS

Formal analysis: TWE

Funding Acquisition: AS

Investigation: TWE

Methodology: TWE, BYY, AC, SK, SMH, AB, AS

Project administration: AS

Resources: AS

Software: TWE, SMH, AB, AS

Supervision: AS

Validation: TWE

Visualization: TWE

Writing – original draft: TWE

Writing – review & editing: TWE, BYY, AC, SK, SMH, AB, AS, TLSB, AN, BAG, JCM

## Data and materials availability

All data were accessed and downloaded between August 2024 and January 2025. All data sources used for this study (A4, ADNI, GS1, GS2, HABS, HABS-HD, OASIS, NACC-SCAN) are publicly available datasets which were accessed following formal applications and data usage agreements. All code written for this manuscript will be shared online (https://github.com/sotiraslab/earnest_nmf_ad_staging). This repository will also include image files for PACs and PTCs as well as code for applying the staging model described herein.

