## Supplementary Materials for "A unified model for staging amyloid and tau pathology in Alzheimer’s disease"

**Supplementary Methods:**

**Dataset-specific PET methods**

A4: [$^{18}$F]-florbetapir images were acquired 50-70 minutes post-injection in 4x5-minute frames. [$^{18}$F]-flortaucipir images were acquired 80-110 minutes post-injection in 6x5-minute frames. Images were provided in multi-frame format. Amyloid-positivity was determined through a combination of automated processing and visual read *(43)*. Amyloid SUVRs were calculated in a composite summary region referenced to the whole cerebellum *(44, 45)*. Individuals were marked as positive if their amyloid SUVR exceeded 1.15. Cases with SUVRs between 1.10-1.15 were determined by a visual read with consensus of two readers.

ADNI: [$^{18}$F]-florbetapir images were acquired 50-70 minutes post-injection in 4x5-minute frames. [$^{18}$F]-florbetaben images were acquired 90-110 minutes post-injection in 4x5-minute frames. [$^{18}$F]-flortaucipir images were acquired 75-105 minutes post-injection in 6x5-minute frames. Images were initially downloaded with frames aligned and average (Co-registered, Averaged). We found some ADNI sites were tau images in this format had intensity artefacts (024, 029, 032, 041, 053, 094, 109, 127, 128); for these, we either downloaded the raw images (if in DICOM or ECAT format) or otherwise used the most processed image with 8mm effective resolution (Coreg, Avg, Std Img and Vox Siz, Uniform Resolution). All amyloid-PET scans were processed by the ADNI PET Core to derive an assessment of amyloid-positivity. Images were processed with FreeSurfer (v7.1.0) using the structural MRI that is temporally closest to the PET image. Regional SUVRs were calculated with the whole cerebellum as a reference region. A global SUVR was calculated in a cortical summary region consisting of frontal, cingulate, lateral parietal, and lateral temporal regions. Amyloid-positivity was determined using SUVR cutoffs of 1.11 (florbetapir) and 1.08 (florbetaben).

GS1/GS2: Amyloid PET scans were captured using [$^{18}$F]-florbetapir , while tau scans were captured with [$^{18}$F]-flortaucipir. Centiloids *(23)* for all subjects were provided, and amyloid-positivity was assessed by a Centiloid cutoff of 24.

HABS: [$^{11}$C]-Pittsburgh Compound B images were acquired 40-60 minutes post-injection in 4x5-minute frames. [$^{18}$F]-flortaucipir images were acquired with two protocols: either 80-100 minutes post-injection in 4x5-minute frames or 75-105 minutes post-injection in 6x5-minute frames. Images were processed with FreeSurfer (v6.0) using the structural MRI that is temporally closest to the PET image. A global amyloid SUVR was calculated in a summary composite consisting of frontal, lateral, and retrosplenial (FLR) regions using cerebellar gray matter as the reference region. Amyloid-positivity was set with an SUVR threshold of 1.28.

HABS-HD: [$^{18}$F]-florbetaben images were acquired 90-110 minutes post-injection in 4x5-minute frames. [$^{18}$F]-PI2620 images were acquired 45-75 minutes post-injection in 6x5-minute frames. Images were processed with FreeSurfer. A global amyloid SUVR was calculated in a composite region matching ADNI2 preprocessing methods, using a whole cerebellum reference region. Amyloid-positivity was defined using a SUVR cutoff of 1.08.

OASIS: [$^{18}$F]-florbetapir images were acquired 50-70 minutes post-injection in 4x5-minute frames. [$^{11}$C]-Pittsburgh Compound B images were acquired 30-60 minutes post-injection in 6x5-minute frames. [$^{18}$F]-flortaucipir images were acquired 75-105 minutes post-injection in 6x5-minute frames. Images were processed using the Pet Unified Pipeline (https://github.com/ysu001/PUP) *(46, 47)* using the T1 image from the same visit. Global amyloid SUVR was calculated in a region consisting of precuneus, prefrontal, gyrus rectus, and lateral temporal cortices using a cerebellum cortex reference region. Amyloid SUVRs were converted to Centiloid and amyloid-positivity with a cutoff of 20.6 for AV45 and 16.4 for PIB.

SCAN: We selected SCAN scans with frame alignment and averaging (Co-registered, Averaged) applied by the SCAN processing team. While multiple active windows were available for some tracers, we only selected the following. [$^{18}$F]-florbetapir images were acquired 50-70 minutes post-injection in 4x5-minute frames. [$^{18}$F]-florbetaben images were acquired 90-110 minutes post-injection in 4x5-minute frames. [$^{11}$C]-Pittsburgh Compound B images were acquired 40-60 minutes post-injection in 4x5-minute frames. [$^{18}$F]-flortaucipir images were acquired 80-100 minutes post-injection in 4x5-minute frames. [$^{18}$F]-PI2620 images were acquired 45-75 minutes post-injection in 6x5-minute frames. We consulted the National Alzheimer's Coordinating Center datasheets for amyloid status (Uniform Data Set 3 field: "AMYLPET").

**MRI and PET processing**

For all PET scans, the T1-weighted MRI scan which was temporally closest to the mean date of amyloid-PET and tau-PET acquisitions was used for preprocessing. Raw T1 DICOMs were converted to NIFTI format, reoriented to right-posterior-inferior (RPI) orientation, and bias-corrected with ANTS N4. DeepMRSeg *(48)* was then applied to skullstrip the image and segment it into 153 regions of interest (ROIs) corresponding to the MUSE algorithm *(38, 49)*. ANTS symmetric diffeomorphic registration was applied to nonlinearly register the brain image to MNI template at 1mm isotropic resolution.

PET images were accessed with varying levels of minimal preprocessing applied by their respective datasets (see Supplementary Material). Raw PET DICOMs were converted to NIFTI format and reoriented to right-posterior-inferior orientation. Multivolume PET images were averaged into a single volume, with FSL MCFLIRT used for motion correction. We applied an iterative smoothing algorithm to enforce a common effective resolution across images (see next section). The smoothed, single volume PET images were then rigidly (6 degrees of freedom) registered to their respective T1 using a mutual information cost function. This registration was applied to move the MUSE ROIs to PET-space, and standardized uptake value ratio (SUVR) images were computed using the cerebellar gray matter as a reference region. The PET-space SUVR images were then warped to MNI space using the combination of the rigid PET-to-TI and nonlinear T1-to-MNI registrations.

Preprocessing outputs for all sessions were manually visually inspected for quality control assurance. Sessions were omitted due to (a) poor alignment of brain mask with the brain, (b) poor alignment of the regional segmentation with the brain, (c) poor alignment of the registered T1 image and the MNI template, and (d) poor coregistration of PET scans with the T1 image.

**PET smoothing**

After frame realignment and averaging but prior to T1-coregistration, we smoothed PET images to achieve a common effective resolution across data sources. Following previous work *(50)*, we used the 3dFWHMx tool from AFNI *(51, 52)* to generate estimates of the PET resolution in the x, y, and z dimensions. The "-2difMAD" and "-automask" options were specified when running 3dFWHMx. From preliminary experiments, we identified that ADNI images labeled to have an effective resolution of 8mm full-width half-maximum (FWHM) had 3dFWHMx estimates around 10mm-isotropic on average (data not shown). As such, we set our target resolution PET to be 10mm-isotropic as measured by 3dFWHMx. To achieve this resolution, we iteratively applied coarser Gaussian smoothing kernels to the original PET image. The smoothing kernel was progressively increased by 0.5mm FHWM in the x, y, and z axes, with a single dimension increased on each iteration. Once the desired smoothness was reached in a given axis (within a 0.5mm FWHM tolerance), the smoothing width for that axis was no longer increased. Iterations proceeded until the image had reached the desired resolution in all directions. In cases which PET images had an initial resolution estimate coarser than 10mm-isoptropic FWHM (due to poorer scanner resolution or smoothing already being applied by the dataset), no additional smoothing was performed.

**Preclinical Alzheimer Cognitive Composite (PACC)**

The PACC is a neuropsychological composite measure developed to detect the earliest cognitive changes occurring in those with preclinical AD *(40)*. Given different neuropsychological assessments available across the datasets we utilized, we applied two previously described formulations of the PACC. For A4, HABS, HABS-HD, and OASIS, we used the original definition, consisting of the free recall score from the Free and Cued Selective Reminding Test *(53)*, the delayed recall score from the Logical Memory IIa of the Wechsler Memory Scale, the score from the Digit Symbol Substitution Test of the Weschler Adult Intelligence Scale-Revised *(54)*, and the MMSE total score. For ADNI, we used the delayed free recall score (question 4) from the Alzheimer's Disease Assessment Scale cognition subscale (expanded version) *(55, 56)*, delayed recall score from the Logical Memory IIa of the Wechsler Memory Scale *(57)*, time to complete the Trail Making Test Part B *(58)*, and the Mini Mental Status Examination (MMSE) score *(39)*. These are the measures recommended for defining PACC in the ADNIMERGE R package (https://adni.bitbucket.io/reference/pacc.html). For GS1, GS2, and SCAN, the PACC was not computed due to a lack of assessments meeting either of these definitions.

PACC scores were computed for each individual if they had at least two contributing assessments present. In case of either PACC definition, the individual contributing assessments were Z-scored. Individuals in the training-NC and validation-NC groups were used to compute the Z-scoring parameters (mean and standard deviation) for the training and validation groups, respectively. Z-scores were scaled such that lower scores indicate more impairment and averaged to compute the PACC. As such, a PACC score of zero indicates the average performance in NC individuals, with negative values indicating progressive impairment.

**NMF training and model selection**

Input data are supplied to the algorithm as a tall $m \times n$ input data matrix $X$, with $m$ being the input feature dimensionality and $n$ being the number of subjects. The objective of NMF is to solve $X \approx WH$, where $W$ is a $m \times k$ factor matrix and $H$ is a $k \times n$ loading matrix. The rank parameter $k$ is user-specified and determines the number of factors estimated, with $k$ typically being much smaller than $m$. The loading matrix contains the factors which capture covariance structures of $X$, and $H$ contains subject-specific coefficients determining how strong each factor contributes to the approximation of an individual subject's data. I.e., all individual observations are approximated as unique linear combination of factors from $W$. Our implementation of NMF *(21)* also constrains $W$ to be orthonormal ($W^T W = I$) and $H$ to be a projection of the data matrix onto the factors ($H \approx W^T X$). These constraints have been shown to engender additional factor sparsity, allowing for a more interpretable interpretation of the data matrix as a sum of parts. Additional details on the formulation of NMF are provided elsewhere *(21, 22, 59, 60)*.

Following previous work, we applied a model selection procedure for selecting the rank parameter $k$ when applying NMF *(15, 22)*. Increasing the rank offers more explanatory power but has the potential to overfit noise in the input data. To balance these notions, we ran NMF multiple times while varying $k$ (from 2-20) and evaluated measures of data fit and factor generalization. Data fit was assessed by plotting the input data reconstruction error

($||X - WH||_{Frobenius}$) at each rank. Factor generalization was assessed using a split-half analysis. For each rank, the data were split into two halves (matched for age, sex, dataset, CDR status, global amyloid burden, and global tau burden) and NMF was applied separately on each half. Factors from each half were matched and the median inner product of matched factors was calculated. The inner product was calculated after rescaling components to have a Euclidean norm of 1, resulting in values between 0 and 1 (with higher values indicating greater similarity). The split-half procedure was repeated 10 times (with results averaged across runs) to account for the variance of data splitting. The final choice of $k$ was achieved by identifying the rank with peak factor generalization which occurs near the rank where data fit begins to plateau.

**W-scoring**

We followed previously described protocols for converting regional PET uptakes to normative deviations in the form of W-scores *(15, 24)*. The general algorithm we used for W-scoring is as follows. The dataset of interest was split into diseased (i.e., ADS) and healthy (i.e., NC) populations. The healthy cohort was split into a training and testing sets (80%/20% split), matched for age, sex, global amyloid burden, and global tau burden. In the training set, a regression model was learned using normative covariates (age and sex) to predict the variable of interest (i.e., PET SUVR in each PAC or PTC). This model was then applied to predict outputs for the test set, and the standard deviation of the model residuals was stored as a measure of the normative deviation in the modeled variable of interest. The model was finally applied to the diseased cohort, and a subject-specific deviation score was calculated as the difference between the observed value and predicted values, divided by the normative deviation (estimated in the test set). The process of splitting the control cohort, estimating the normative uptake deviation, and calculating the deviation score was repeated 200 times to account for the variability of data splitting. The final W-score for each subject was calculated as the average of the 200 deviation scores.

We calculated W-scores for PET uptakes in PACs and PTCs, the amyloid and tau factors estimated with NMF. W-score calculation was done separately for training and validation sets, using the ADS groups as the diseased population and the NC groups as the healthy population. Scores were also calculated separately for the different subsets of the validation set to ensure the calculated values were tracer specific (validation-A: FBB/FTP, validation-B: FBB/PI2620, validation-C: PIB/FTP). Following prior work, W-scores were thresholded at 2.5 to determine the binary presence or absence of pathology within PACs and PTCs *(15, 24)*.

**Bootstrapped staging algorithm**

We employed our previously described approach for converting distributions of regional pathology in PACs and PTCs into a staging model *(15)*. Like other methods for staging in neurodegenerative disease, this approach infers the temporal ordering of regions in a pathological cascade through observed cross-sectional rates of pathology. Namely, regions with higher observed frequencies of pathology are assumed to develop pathology earlier. The present method extends this notion by aiming to identify regions that are temporally similar by determining pairs of regions which have statistically similar rates of pathology. The input to the model is a table containing a binary assessment of pathology (present/absent) for all regions and all subjects. Bootstrapping is then used to resample the dataset and estimate null models for pairwise differences in pathology rates between regions. In each bootstrap sample, the null hypothesis (regions have equal rates of pathology) is modeled by subtracting the observed regional frequency of pathology (calculated in the non-bootstrapped data) from that of the bootstrap sample. This correction, which may result in negative values, and ensures that on average (across bootstrap iterations) the frequency of elevated for pathology is centered at zero and equal for all regions. In this null data, pairwise differences in regional frequencies are calculated. This process is repeated 5,000 to model a null distribution. Finally, a p-value for each pair of regions is calculated by counting the number of times the bootstrapped difference in pathology elevation is greater or equal than the observed difference in the full dataset. A false discovery rate (FDR) correction is applied to these p-values to correct for multiple comparisons *(42)*. Regions are then grouped into stages by grouping together regions which have non-significant ($p>0.05$) p-values, with the order determined by the total positivity rate.

**Permutation test for longitudinal staging**

For all individuals with longitudinal scanning, we assembled a table containing the assigned stage at the first and last visit. We then randomly permuted (5,000 permutations) the order of first and last scans based on the outcome of a Bernoulli trial ($p=0.5$) for each individual. A p-value was calculated by counting the number of permutations where the proportion of individuals with expected progression (i.e., the final stage is equal to or more advanced than the first stage) was greater or equal to the observed data. For these analyses, NS scans were included but counted as non-expected progression.

**Supplementary Figures:**

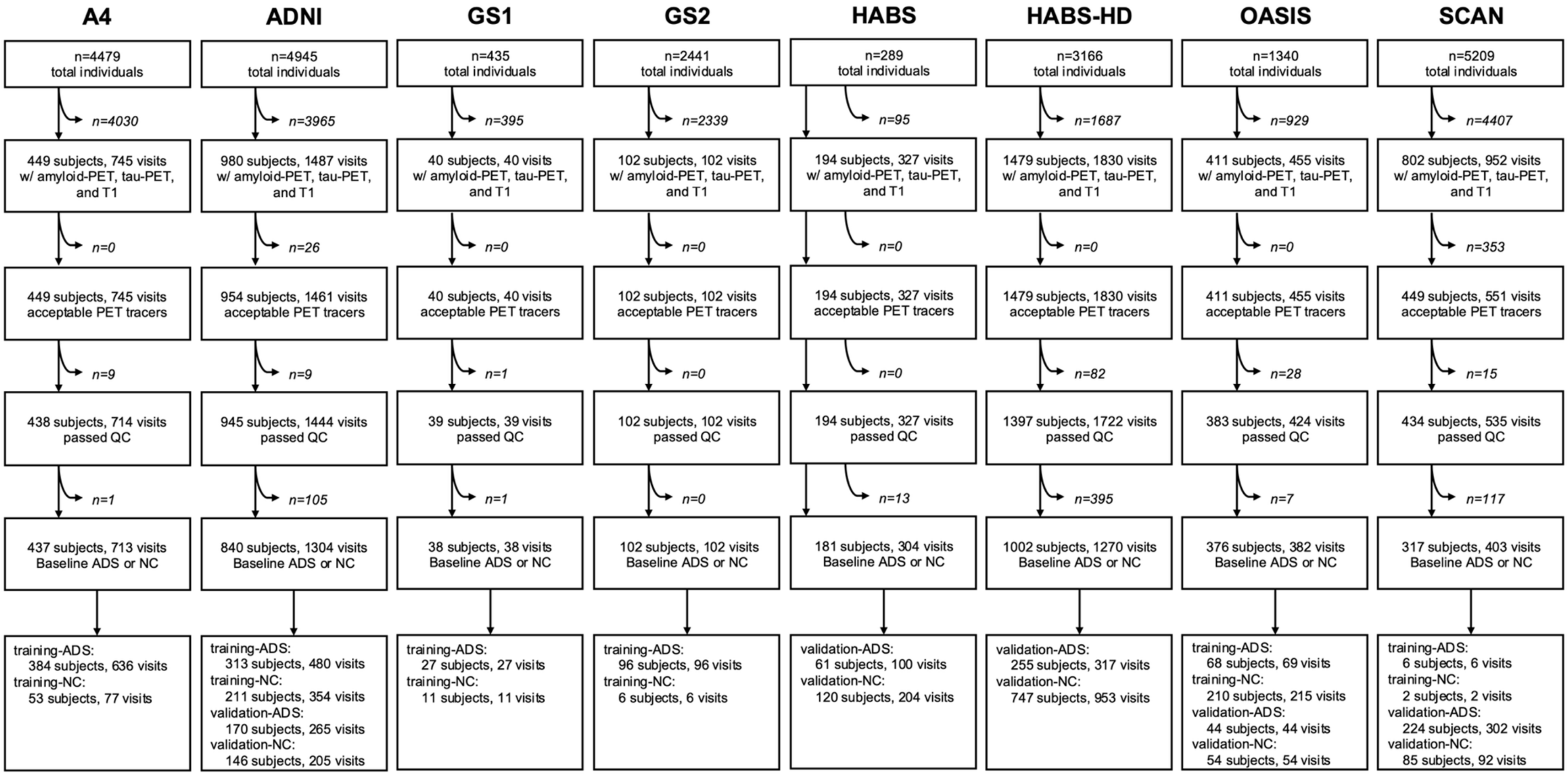


**Supplementary Figure 1. Flowchart of participant inclusion and exclusion.** Separate flow charts are shown for each source dataset. Curved arrows with italicized text represent counts of excluded individuals. The top cell represents the total number of individuals with any imaging studies done. The final row shows how many subjects were included in the final analysis. "Visits" refers to unique sessions where a subject has amyloid-PET and tau-PET. "Acceptable PET tracers" refers to the tracer pairs required for the training set (FBP/FTP) or the validation set (FBB/FTP; FBB/P26; PIB/FTP). ADS=Alzheimer's Disease Spectrum, NC=normative control, QC=quality control.

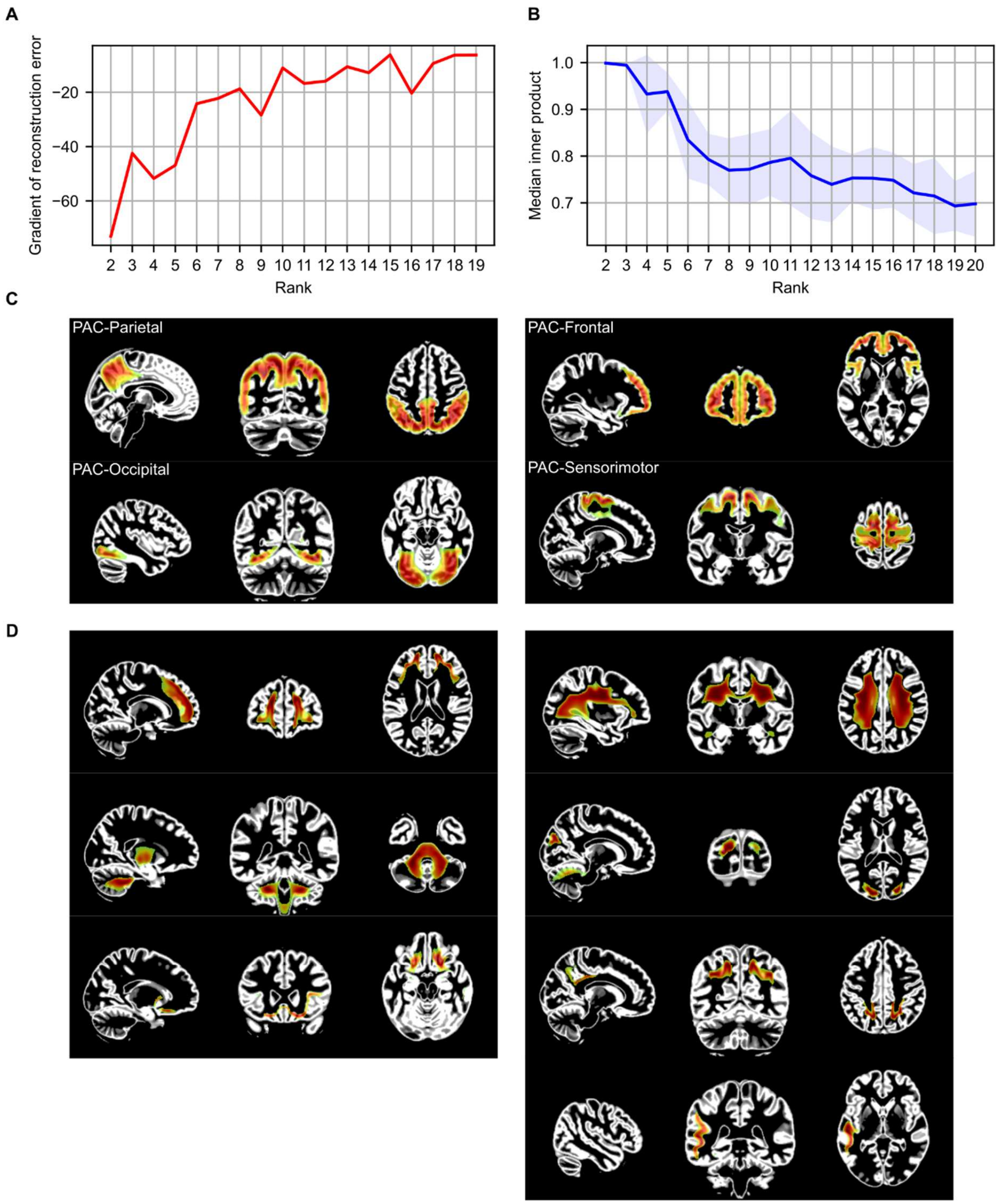


**Supplementary Figure 2. NMF model selection for the estimation of amyloid factors (PACs).** NMF was applied to amyloid-PET volumes of the training data (n=1,387) at different ranks (2-20). **(A)** Gradient of reconstruction error of NMF solutions, showing a plateau between 8-12 factors. **(B)** Similarity of factors estimated separately with split half resampling. In each iteration, factors estimated in different subsets are matched and the inner product of pairs of factors are recorded and summarized with a median. The blue line shows the average of median inner products over 10 repeats with different random seeds, while the blue area shows the standard deviation. Local peaks in reproducibility were observed at 2, 5, 11, and 14 factors. The 11-factor model was chosen for further analysis. **(C)** Factors from the rank 11 solution which overlapped with cortical gray matter and were included in the main analysis. **(D)** Omitted factors from the rank 11 solution, capturing primarily off-target binding. For (C) and (D), factor distributions are shown in color over a gray matter partial volume estimate map in MNI space. NMF=non-negative matrix factorization; PAC=Pattern of Amyloid Covariance.

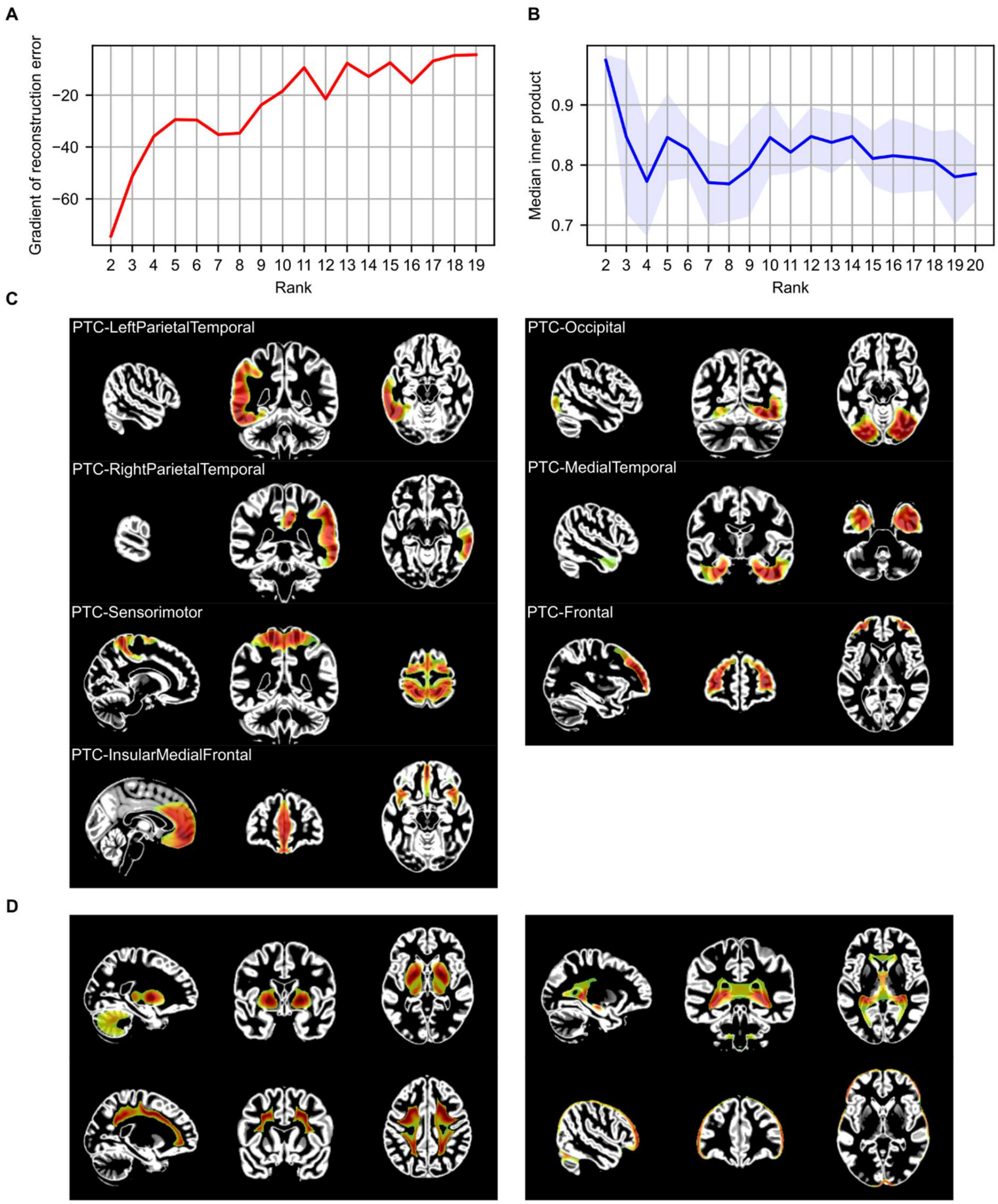


**Supplementary Figure 3. NMF model selection for the estimation of tau factors (PTCs).** NMF was applied to tau-PET volumes of the training data (n=1,387) at different ranks (2-20). **(A)** Gradient of reconstruction error of NMF solutions, showing a plateau between 10-13 factors. **(B)** Similarity of factors estimated separately with split half resampling. In each iteration, factors estimated in different subsets are matched and the inner product of pairs of factors are recorded and summarized with a median. The blue line shows the average of median inner products over

10 repeats with different random seeds, while the blue area shows the standard deviation. Local peaks in reproducibility were observed at 2, 5, 10, 12, and 14 factors. The 12-factor model was chosen for further analysis. **(C)** Factors from the rank 12 solution which overlapped with cortical gray matter and were included in the main analysis. **(D)** Omitted factors from the rank 11 solution, capturing primarily off-target binding. For (C) and (D), factor distributions are shown in color over a gray matter partial volume estimate map in MNI space. NMF=non-negative matrix factorization; PTC=Pattern of Tau Covariance.

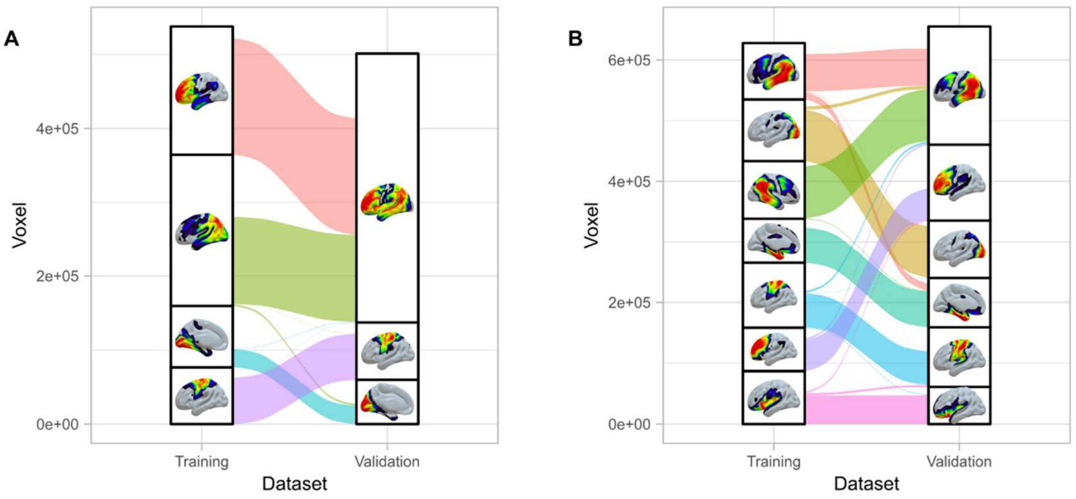


**Supplementary Figure 4. Alluvial plots showing similarity of NMF solutions for the training and testing data.** **(A)** Comparison of factors for amyloid. **(B)** Comparison of factors for tau. In each panel, the left stack shows the training factors while the right stack shows the validation factors. Voxels were assigned to factors using a winner-takes-all procedure after filtering the factors to remove areas with minimal loadings. Each box represents a single factor with height proportional to the number of voxels assigned to it. Colored flows indicate how individual voxels are assigned under the training and validation factorizations. Flows are not shown for voxels which were not contained in one factorization (due to being filtered out or being assigned to a non-cortical gray matter factor). Areas with missing flows indicate portions of the factor which do not overlap spatially with any factor in the other dataset. Both amyloid and tau show indications of hierarchical splitting and joining of NMF factors across training and validation solutions. E.g., in (A) the first validation factor is primarily composed of the first two (green/red) training factors. Likewise, in (B), the first validation factor is primarily composed of the first and third (green/red) training factors.

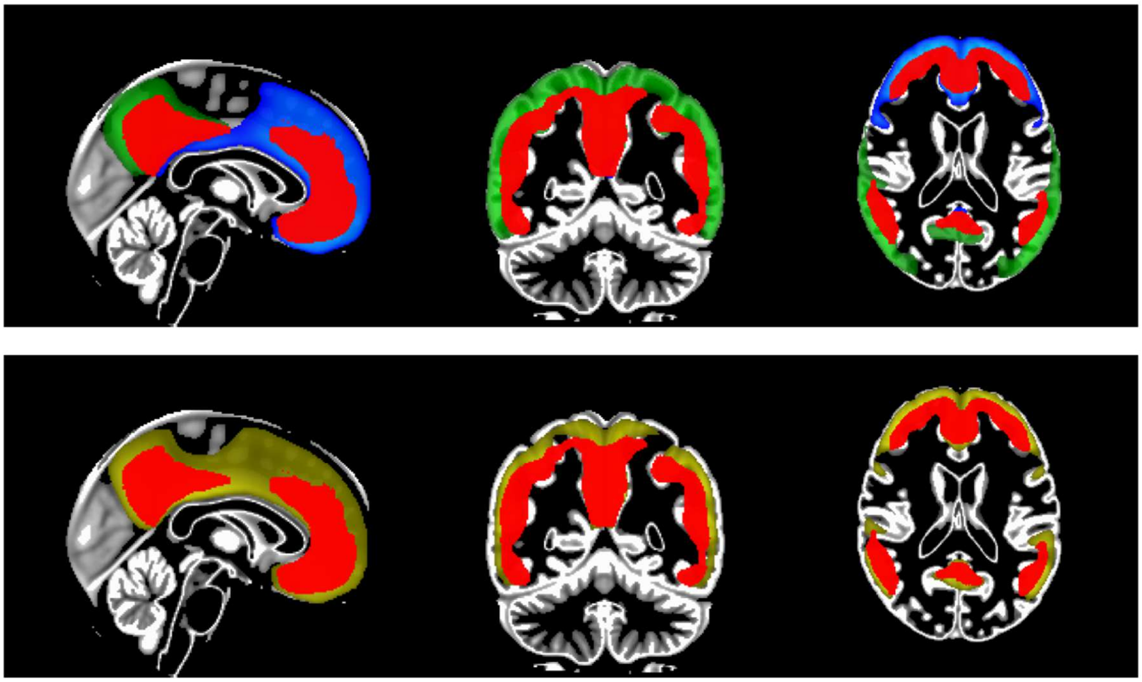

**Supplementary Figure 5. Comparison of frontal and parietal amyloid factors with the Centiloid mask. The top panel shows the overlap of PAC-Frontal (blue) and PAC-Parietal (green) with the Centiloid mask (red). The bottom panel shows the overlap of a factor estimated from the validation dataset (yellow).**

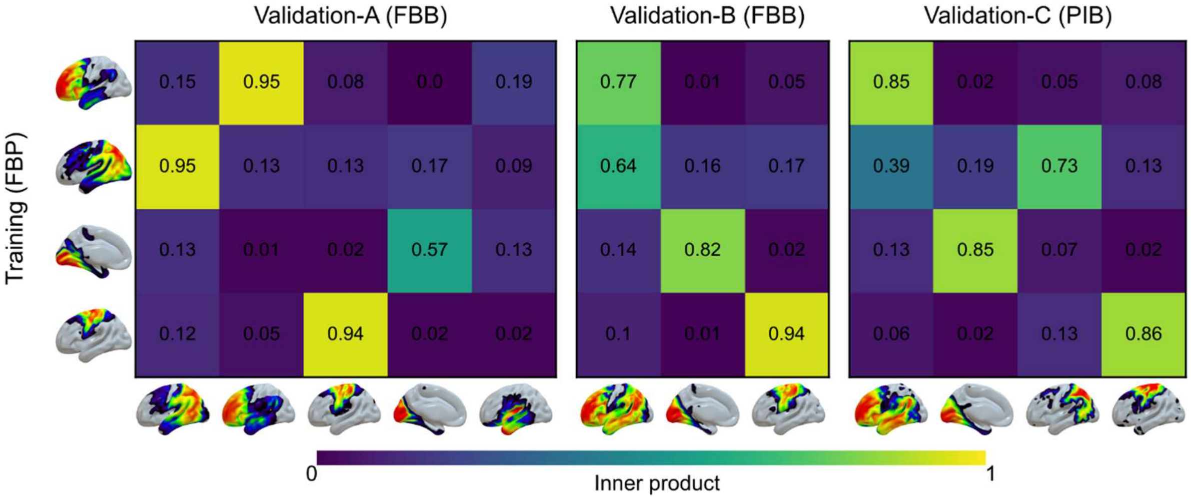


**Supplementary Figure 6. Comparison of amyloid factors with those estimated in subsets of the validation data.** NMF was rerun on the subsets of the validation dataset to test model reproducibility under different PET tracers. As in the training set, NMF was run at rank 11 and factors not highlighting cortical gray matter areas were removed. Heatmaps show the inner product between matched factors. From left to right, subplots show the similarity for validation-A (FBB; n=486), validation-B (FBB; 1,097), and validation-C (PIB; 323). FBB=florbetaben; FBP=florbetapir; NMF=non-negative matrix factorization; PIB=Pittsburgh Compound B.

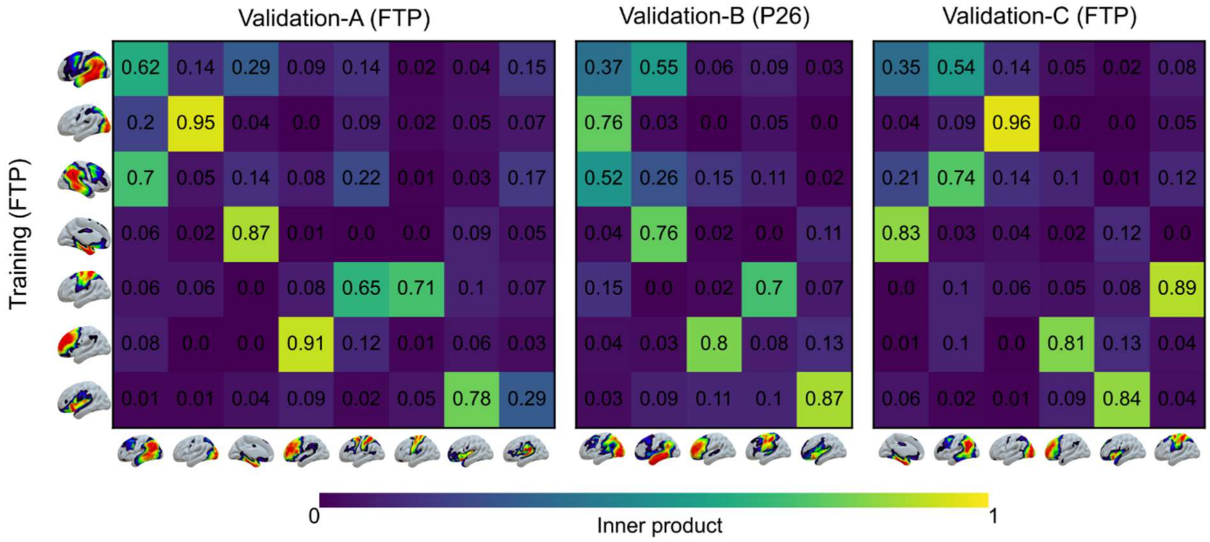


**Supplementary Figure 7. Comparison of tau factors with those estimated in subsets of the validation data.** NMF was rerun on the subsets of the validation dataset to test model reproducibility under different PET tracers. As in the training set, NMF was run at rank 12 and factors not highlighting cortical gray matter areas were removed. Heatmaps show the inner product between matched factors. From left to right, subplots show the similarity for validation-A (FTP; n=486), validation-B (P26; 1,097), and validation-C (FTP; 323). FTP=flortaucipir; NMF=non-negative matrix factorization; P26=PI-2620.

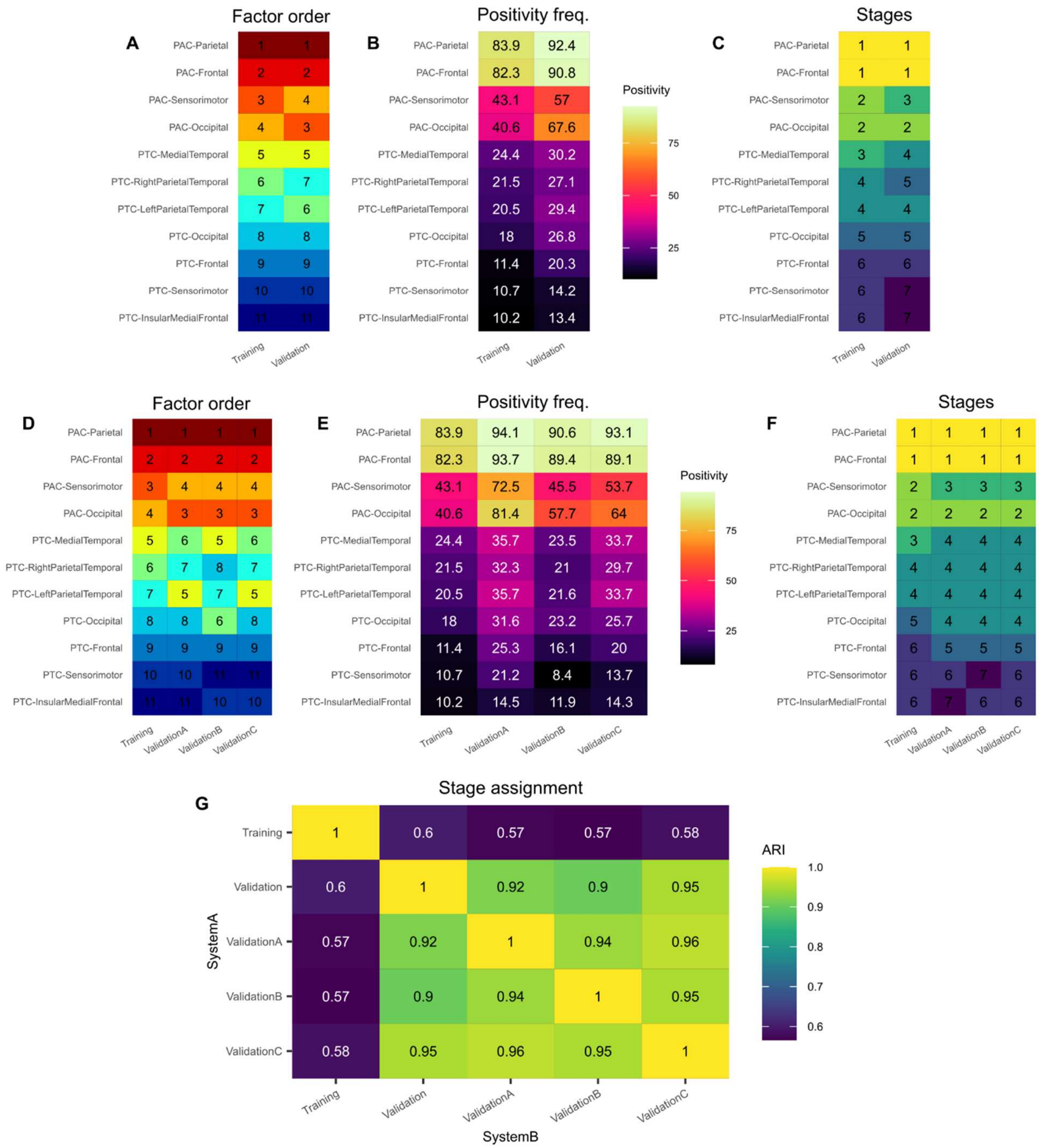


**Supplementary Figure 8. Replication of staging analyses using validation data.** The distribution of W-scores within PACs and PTCs in the validation set was used to replicate the creation of the staging model derived from training data. All references to "Training" in this figure refer to the model presented in the main text **(A)** Ordering of regions based on the frequency of suprathreshold W-scores in training-ADS (n=894) and validation-ADS data (n=754). **(B)** Rate of positivity in each PAC and PTC. **(C)** The grouping of PACs and PTCs into stages based on models developed in training and replication data. **(D-F)** Same as (A-C), but showing comparisons with the fixed tracer subsets of the validation dataset (validation-A: n=486; validation-B: n=1,097; validation-C: n=323). **(G)** The training model and all validation models were used to assign labels to all training-ADS and validation-ADS subjects. The heatmap shows the adjusted rand index (ARI) for stage assignments of the same subjects under different staging models.

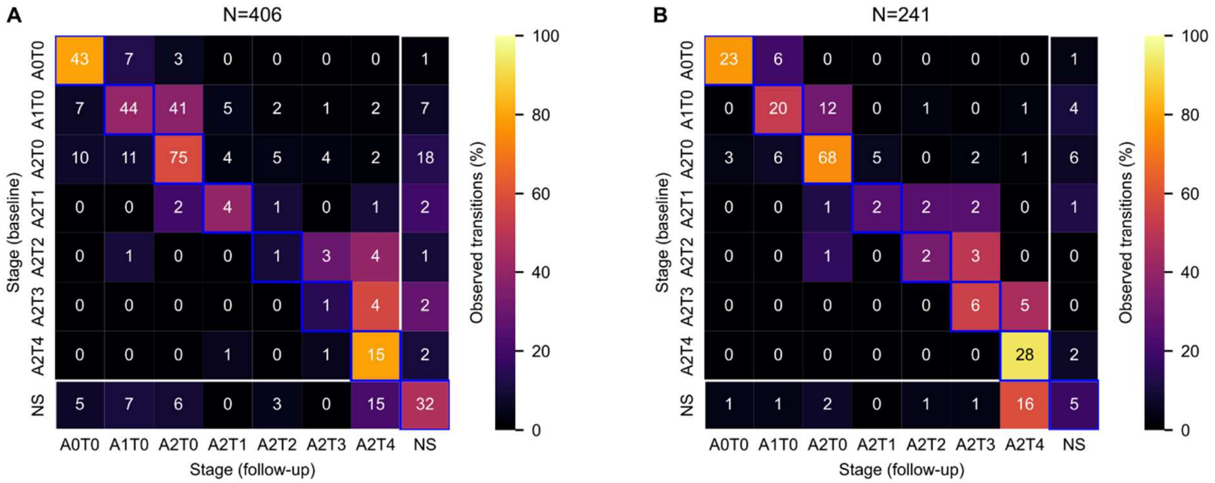


**Supplementary Figure 9. Longitudinal transitions of stages.** **(A)** Longitudinal stage transitions for training-ADS. **(B)** Longitudinal stage transitions for validation-ADS. Both panels show the cross tabulation of each individual's stage at baseline (rows) versus their stage at their final follow-up visit (columns). Text within each cell shows the count of individuals. Cell colors show the percentage of individuals in each cell normalized to the sum across the row. The blue boxes highlight individuals who do not change stage from baseline to follow-up. Excluding the last row and column (non-stageable presentations), cells on (no change) or above (advance in stage) the diagonal represent cases which conform to the staging model, while observations below the diagonal (reversion in stage) indicate deviations from the expected staging model.

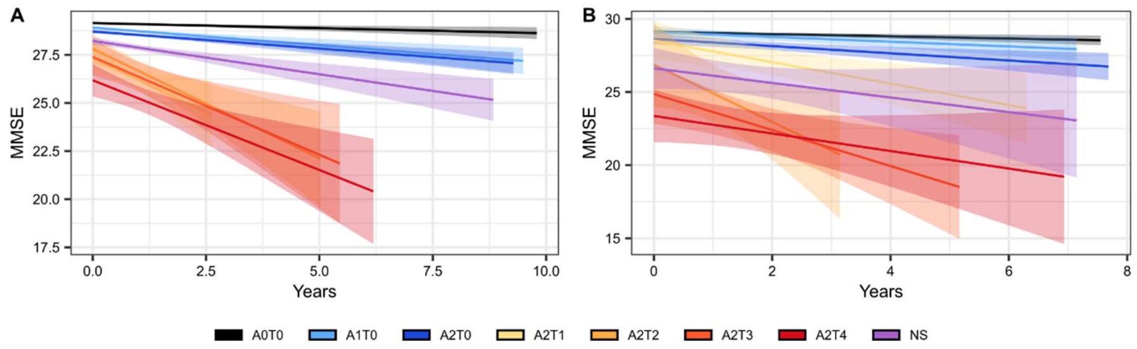


**Supplementary Figure 10. Mixed effect models comparing longitudinal MMSE scores across disease stages.** (**A**) Estimated fit lines for the model fit to training data. (**B**) Estimated fit lines for the model fit to validation data.

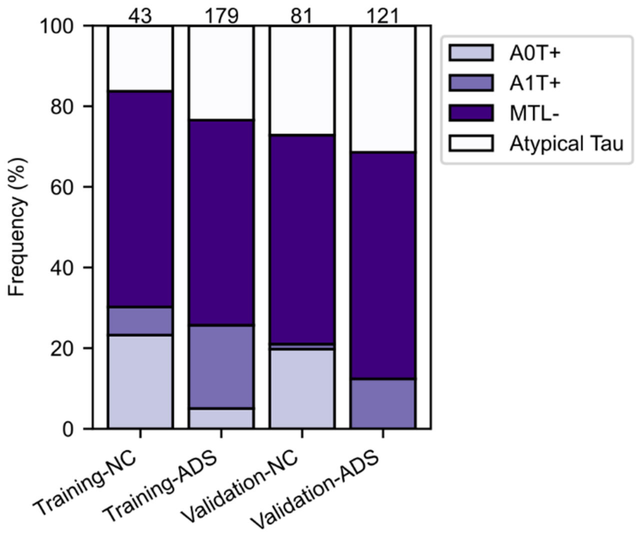


**Supplementary Figure 11. Boxplot showing the further characterization of stages for those labeled as non-stageable (NS).** Counts for each group are printed at the top of each bar. A0T+: tau pathology without any amyloid, A1T+: tau and late stage amyloid without early amyloid, MTL-: both stages of amyloid and some tau without PTC-MedialTemporal positivity, Atypical Tau: all other NS cases.

**Supplementary Tables:**

| Biomarker | FreeSurfer region | MUSE region |
|---|---|---|
| Amyloid | Caudal anterior cingulate | Middle cingulate gyrus (MCgG) |
| Amyloid | Caudal middle frontal | Middle frontal gyrus (MFG) |
| Amyloid | Frontal pole | Frontal pole (FRP) |
| Amyloid | Inferior parietal | Angular gyrus (AnG) |
| Amyloid | Inferior temporal gyrus | Inferior temporal gyrus (ITG) |
| Amyloid | Isthmus cingulate | Posterior cingulate gyrus (PCgG) |
| Amyloid | Medial orbitofrontal | Subcallosal area (SCA) |
| Amyloid | Middle temporal | Middle temporal gyrus (MTG) |
| Amyloid | Pars opercularis | Opercular part of the inferior frontal gyrus (OpIFG) |
| Amyloid | Pars orbitalis | Orbital part of the inferior frontal gyrus (OrIFG) |
| Amyloid | Pars traingularis | Triangular part of the inferior frontal gyrus (TrIFG) |
| Amyloid | Posterior cingulate | Middle cingulate gyrus (MCgG) |
| Amyloid | Precuneus | Precuneus (PCu) |
| Amyloid | Rostral anterior cingulate | Anterior cingulate gyrus (ACgG) |
| Amyloid | Rostral middle frontal | Anterior orbital gyrus (AOrG), middle frontal gyrus (MFG) |
| Amyloid | Superior frontal | Superior frontal gyrus medial segment (MSFG), superior frontal gyrus (SFG), supplementary motor cortex (SMC) |
| Amyloid | Superior parietal | Superior parietal lobule (SPL) |
| Amyloid | Superior temporal | Planum polare (PP), planum temporale (PT), superior temporal gyrus (STG) |
| Amyloid | Supramarginal | Parietal operculum (PO), supplementary motor cortex (SMC) |
| Tau | Amygdala | Amygdala |
| Tau | Entorhinal | Entorhinal area (Ent) |
| Tau | Fusiform | Fusiform gyrus (FuG), occipital fusiform gyrus (OFuG) |
| Tau | Inferior temporal | Inferior temporal gyrus (ITG) |
| Tau | Middle temporal | Middle temporal gyrus (MTG) |

**Supplementary Table 1. Regions of interest used for global amyloid and tau burden.** FreeSurfer regions used to define composites were taken from the ADNI PET Core methods and translated to MUSE regions of interest *(38)* for our analysis. Both left and right hemisphere regions were used for all listed areas. Some MUSE regions appear multiple times (e.g., middle frontal gyrus) as they overlap with multiple FreeSurfer regions. Such regions were not counted multiple times for the purpose of creating the volume-weighted average SUVR.

| Comparison | Difference | Lower | Upper | p-value | Significance |
|---|---|---|---|---|---|
| **A1T0-A0T0** | 0.27 | -0.06 | 0.59 | 0.206 | |
| **A2T0-A0T0** | 0.37 | 0.05 | 0.68 | 0.01 | ** |
| **A2T1-A0T0** | 1 | 0.1 | 1.89 | 0.017 | * |
| **A2T2-A0T0** | 1.7 | 0.93 | 2.46 | <0.001 | *** |
| **A2T3-A0T0** | 2.45 | 1.42 | 3.48 | <0.001 | *** |
| **A2T4-A0T0** | 2.87 | 2.35 | 3.39 | <0.001 | *** |
| **NS-A0T0** | 0.66 | 0.33 | 0.98 | <0.001 | *** |
| **A2T0-A1T0** | 0.1 | -0.27 | 0.47 | 0.991 | |
| **A2T1-A1T0** | 0.73 | -0.19 | 1.65 | 0.232 | |
| **A2T2-A1T0** | 1.43 | 0.64 | 2.22 | <0.001 | *** |
| **A2T3-A1T0** | 2.18 | 1.13 | 3.23 | <0.001 | *** |
| **A2T4-A1T0** | 2.6 | 2.04 | 3.16 | <0.001 | *** |
| **NS -A1T0** | 0.39 | 0.01 | 0.77 | 0.044 | * |
| **A2T1-A2T0** | 0.63 | -0.28 | 1.54 | 0.421 | |
| **A2T2-A2T0** | 1.33 | 0.54 | 2.11 | <0.001 | *** |
| **A2T3-A2T0** | 2.08 | 1.03 | 3.13 | <0.001 | *** |
| **A2T4-A2T0** | 2.5 | 1.95 | 3.06 | <0.001 | *** |
| **NS -A2T0** | 0.29 | -0.09 | 0.66 | 0.282 | |
| **A2T2-A2T1** | 0.7 | -0.45 | 1.85 | 0.586 | |
| **A2T3-A2T1** | 1.45 | 0.11 | 2.79 | 0.023 | * |
| **A2T4-A2T1** | 1.87 | 0.87 | 2.88 | <0.001 | *** |
| **NS -A2T1** | -0.34 | -1.26 | 0.58 | 0.951 | |
| **A2T3-A2T2** | 0.75 | -0.51 | 2.01 | 0.611 | |
| **A2T4-A2T2** | 1.17 | 0.28 | 2.06 | 0.002 | ** |
| **NS -A2T2** | -1.04 | -1.83 | -0.25 | 0.002 | ** |
| **A2T4-A2T3** | 0.42 | -0.7 | 1.55 | 0.948 | |
| **NS -A2T3** | -1.79 | -2.84 | -0.74 | <0.001 | *** |
| **NS-A2T4** | -2.21 | -2.78 | -1.65 | <0.001 | *** |

**Supplementary Table 2. Tukey post-hoc differences for the ANOVA comparing CDR-SB scores across stages in the training set.** Lower and Upper contain the lower and upper bounds of the 95% confidence interval of the difference in means.

| Comparison | Difference | Lower | Upper | p-value | Significance |
|---|---|---|---|---|---|
| **A1T0-A0T0** | -0.31 | -0.79 | 0.17 | 0.514 | |
| **A2T0-A0T0** | -0.71 | -1.19 | -0.23 | <0.001 | *** |
| **A2T1-A0T0** | -2.38 | -3.76 | -0.99 | <0.001 | *** |
| **A2T2-A0T0** | -2.54 | -3.71 | -1.36 | <0.001 | *** |
| **A2T3-A0T0** | -2.73 | -4.26 | -1.19 | <0.001 | *** |
| **A2T4-A0T0** | -3.95 | -4.78 | -3.13 | <0.001 | *** |
| **NS-A0T0** | -1.1 | -1.61 | -0.59 | <0.001 | *** |
| **A2T0-A1T0** | -0.41 | -0.95 | 0.14 | 0.311 | |
| **A2T1-A1T0** | -2.07 | -3.48 | -0.66 | <0.001 | *** |
| **A2T2-A1T0** | -2.23 | -3.43 | -1.03 | <0.001 | *** |
| **A2T3-A1T0** | -2.42 | -3.98 | -0.86 | <0.001 | *** |
| **A2T4-A1T0** | -3.64 | -4.51 | -2.78 | <0.001 | *** |
| **NS -A1T0** | -0.79 | -1.36 | -0.22 | 0.001 | *** |
| **A2T1-A2T0** | -1.66 | -3.07 | -0.25 | 0.009 | ** |
| **A2T2-A2T0** | -1.82 | -3.02 | -0.62 | <0.001 | *** |
| **A2T3-A2T0** | -2.01 | -3.57 | -0.45 | 0.002 | ** |
| **A2T4-A2T0** | -3.24 | -4.1 | -2.38 | <0.001 | *** |
| **NS -A2T0** | -0.39 | -0.96 | 0.18 | 0.439 | |
| **A2T2-A2T1** | -0.16 | -1.93 | 1.61 | 1 | |
| **A2T3-A2T1** | -0.35 | -2.38 | 1.68 | 1 | |
| **A2T4-A2T1** | -1.58 | -3.14 | -0.01 | 0.046 | * |
| **NS -A2T1** | 1.27 | -0.15 | 2.7 | 0.118 | |
| **A2T3-A2T2** | -0.19 | -2.08 | 1.7 | 1 | |
| **A2T4-A2T2** | -1.42 | -2.79 | -0.04 | 0.038 | * |
| **NS -A2T2** | 1.44 | 0.22 | 2.65 | 0.008 | ** |
| **A2T4-A2T3** | -1.23 | -2.92 | 0.47 | 0.355 | |
| **NS -A2T3** | 1.62 | 0.06 | 3.19 | 0.036 | * |
| **NS-A2T4** | 2.85 | 1.97 | 3.73 | <0.001 | *** |

**Supplementary Table 3. Tukey post-hoc differences for the ANOVA comparing MMSE scores across stages in the training set.** Lower and Upper contain the lower and upper bounds of the 95% confidence interval of the difference in means.

| Comparison | Difference | Lower | Upper | p-value | Significance |
|---|---|---|---|---|---|
| **A1T0-A0T0** | 0.27 | -0.05 | 0.59 | 0.171 | |
| **A2T0-A0T0** | 0.66 | 0.41 | 0.91 | <0.001 | *** |
| **A2T1-A0T0** | 1.52 | 0.62 | 2.43 | <0.001 | *** |
| **A2T2-A0T0** | 1.97 | 1.04 | 2.9 | <0.001 | *** |
| **A2T3-A0T0** | 3.23 | 2.49 | 3.97 | <0.001 | *** |
| **A2T4-A0T0** | 4.68 | 4.28 | 5.09 | <0.001 | *** |
| **NS-A0T0** | 0.99 | 0.69 | 1.3 | <0.001 | *** |
| **A2T0-A1T0** | 0.39 | 0.02 | 0.76 | 0.033 | * |
| **A2T1-A1T0** | 1.25 | 0.31 | 2.2 | 0.002 | ** |
| **A2T2-A1T0** | 1.7 | 0.73 | 2.67 | <0.001 | *** |
| **A2T3-A1T0** | 2.96 | 2.17 | 3.75 | <0.001 | *** |
| **A2T4-A1T0** | 4.41 | 3.93 | 4.9 | <0.001 | *** |
| **NS -A1T0** | 0.72 | 0.32 | 1.13 | <0.001 | *** |
| **A2T1-A2T0** | 0.86 | -0.06 | 1.79 | 0.087 | |
| **A2T2-A2T0** | 1.31 | 0.36 | 2.26 | 0.001 | *** |
| **A2T3-A2T0** | 2.57 | 1.8 | 3.33 | <0.001 | *** |
| **A2T4-A2T0** | 4.02 | 3.58 | 4.47 | <0.001 | *** |
| **NS -A2T0** | 0.33 | -0.02 | 0.69 | 0.084 | |
| **A2T2-A2T1** | 0.44 | -0.84 | 1.73 | 0.967 | |
| **A2T3-A2T1** | 1.7 | 0.55 | 2.86 | <0.001 | *** |
| **A2T4-A2T1** | 3.16 | 2.18 | 4.14 | <0.001 | *** |
| **NS -A2T1** | -0.53 | -1.47 | 0.41 | 0.681 | |
| **A2T3-A2T2** | 1.26 | 0.08 | 2.44 | 0.026 | * |
| **A2T4-A2T2** | 2.72 | 1.72 | 3.72 | <0.001 | *** |
| **NS -A2T2** | -0.97 | -1.94 | -0.01 | 0.046 | * |
| **A2T4-A2T3** | 1.46 | 0.63 | 2.29 | <0.001 | *** |
| **NS -A2T3** | -2.23 | -3.02 | -1.45 | <0.001 | *** |
| **NS-A2T4** | -3.69 | -4.17 | -3.21 | <0.001 | *** |

**Supplementary Table 4. Tukey post-hoc differences for the ANOVA comparing CDR-SB scores across stages in the validation set.** Lower and Upper contain the lower and upper bounds of the 95% confidence interval of the difference in means.

| Comparison | Difference | Lower | Upper | p-value | Significance |
|---|---|---|---|---|---|
| **A1T0-A0T0** | -0.24 | -0.84 | 0.37 | 0.937 | |
| **A2T0-A0T0** | -0.43 | -0.94 | 0.08 | 0.172 | |
| **A2T1-A0T0** | 0.03 | -1.83 | 1.9 | 1 | |
| **A2T2-A0T0** | -2.09 | -4.41 | 0.22 | 0.111 | |
| **A2T3-A0T0** | -3.36 | -5.05 | -1.67 | <0.001 | *** |
| **A2T4-A0T0** | -4.94 | -5.98 | -3.91 | <0.001 | *** |
| **NS-A0T0** | -1.19 | -1.8 | -0.58 | <0.001 | *** |
| **A2T0-A1T0** | -0.2 | -0.92 | 0.53 | 0.992 | |
| **A2T1-A1T0** | 0.27 | -1.66 | 2.2 | 1 | |
| **A2T2-A1T0** | -1.86 | -4.23 | 0.51 | 0.254 | |
| **A2T3-A1T0** | -3.13 | -4.9 | -1.36 | <0.001 | *** |
| **A2T4-A1T0** | -4.71 | -5.86 | -3.55 | <0.001 | *** |
| **NS -A1T0** | -0.96 | -1.76 | -0.16 | 0.007 | ** |
| **A2T1-A2T0** | 0.47 | -1.44 | 2.37 | 0.996 | |
| **A2T2-A2T0** | -1.66 | -4.01 | 0.69 | 0.386 | |
| **A2T3-A2T0** | -2.93 | -4.67 | -1.19 | <0.001 | *** |
| **A2T4-A2T0** | -4.51 | -5.62 | -3.4 | <0.001 | *** |
| **NS -A2T0** | -0.76 | -1.49 | -0.03 | 0.034 | * |
| **A2T2-A2T1** | -2.13 | -5.08 | 0.83 | 0.362 | |
| **A2T3-A2T1** | -3.39 | -5.89 | -0.9 | 0.001 | ** |
| **A2T4-A2T1** | -4.98 | -7.08 | -2.87 | <0.001 | *** |
| **NS -A2T1** | -1.23 | -3.16 | 0.71 | 0.534 | |
| **A2T3-A2T2** | -1.27 | -4.12 | 1.58 | 0.879 | |
| **A2T4-A2T2** | -2.85 | -5.37 | -0.33 | 0.014 | * |
| **NS -A2T2** | 0.9 | -1.47 | 3.27 | 0.945 | |
| **A2T4-A2T3** | -1.58 | -3.54 | 0.38 | 0.218 | |
| **NS -A2T3** | 2.17 | 0.4 | 3.94 | 0.005 | ** |
| **NS-A2T4** | 3.75 | 2.59 | 4.91 | <0.001 | *** |

**Supplementary Table 5. Tukey post-hoc differences for the ANOVA comparing MMSE scores across stages in the validation set.** Lower and Upper contain the lower and upper bounds of the 95% confidence interval of the difference in means.

| Contrast | Difference | SE | DF | T ratio | p-value | Annotation |
|---|---|---|---|---|---|---|
| **A0T0 - A1T0** | -0.374 | 0.139 | 1214.441 | -2.695 | 0.125 | |
| **A0T0 - A2T0** | -0.505 | 0.138 | 1216.909 | -3.654 | 0.007 | ** |
| **A0T0 - A2T1** | -2.205 | 0.413 | 1208.212 | -5.344 | <0.001 | *** |
| **A0T0 - A2T2** | -2.408 | 0.33 | 1214.578 | -7.306 | <0.001 | *** |
| **A0T0 - A2T3** | -3.034 | 0.446 | 1304.646 | -6.806 | <0.001 | *** |
| **A0T0 - A2T4** | -4.15 | 0.263 | 1243.41 | -15.753 | <0.001 | *** |
| **A0T0 - Atypical** | -1.114 | 0.149 | 1201.295 | -7.485 | <0.001 | *** |
| **A1T0 - A2T0** | -0.132 | 0.156 | 1215.789 | -0.846 | 0.99 | |
| **A1T0 - A2T1** | -1.831 | 0.418 | 1205.739 | -4.376 | <0.001 | *** |
| **A1T0 - A2T2** | -2.034 | 0.337 | 1212.636 | -6.035 | <0.001 | *** |
| **A1T0 - A2T3** | -2.661 | 0.451 | 1301.06 | -5.894 | <0.001 | *** |
| **A1T0 - A2T4** | -3.776 | 0.273 | 1242.469 | -13.828 | <0.001 | *** |
| **A1T0 - Atypical** | -0.741 | 0.166 | 1209.004 | -4.474 | <0.001 | *** |
| **A2T0 - A2T1** | -1.7 | 0.417 | 1199.549 | -4.071 | 0.001 | *** |
| **A2T0 - A2T2** | -1.902 | 0.336 | 1204.418 | -5.663 | <0.001 | *** |
| **A2T0 - A2T3** | -2.529 | 0.451 | 1297.028 | -5.611 | <0.001 | *** |
| **A2T0 - A2T4** | -3.644 | 0.272 | 1234.529 | -13.394 | <0.001 | *** |
| **A2T0 - Atypical** | -0.609 | 0.164 | 1195.034 | -3.705 | 0.005 | ** |
| **A2T1 - A2T2** | -0.203 | 0.513 | 1199.908 | -0.396 | 1 | |
| **A2T1 - A2T3** | -0.829 | 0.594 | 1253.48 | -1.395 | 0.86 | |
| **A2T1 - A2T4** | -1.944 | 0.474 | 1211.778 | -4.104 | 0.001 | *** |
| **A2T1 - Atypical** | 1.091 | 0.422 | 1201.302 | 2.587 | 0.161 | |
| **A2T2 - A2T3** | -0.627 | 0.54 | 1267.768 | -1.159 | 0.943 | |
| **A2T2 - A2T4** | -1.742 | 0.404 | 1218.553 | -4.314 | <0.001 | *** |
| **A2T2 - Atypical** | 1.294 | 0.341 | 1205.43 | 3.795 | 0.004 | ** |
| **A2T3 - A2T4** | -1.115 | 0.503 | 1288.378 | -2.215 | 0.343 | |
| **A2T3 - Atypical** | 1.92 | 0.454 | 1296.16 | 4.226 | 0.001 | *** |
| **A2T4 - Atypical** | 3.035 | 0.278 | 1232.031 | 10.923 | <0.001 | *** |

**Supplementary Table 6. Post-hoc testing comparing estimated marginal means of the mixed effect model predicting longitudinal CDR-SB scores in the training set.**

| Contrast | Difference | SE | DF | T ratio | p-value | Annotation |
|---|---|---|---|---|---|---|
| **A0T0 - A1T0** | 0.377 | 0.175 | 1165.189 | 2.153 | 0.382 | |
| **A0T0 - A2T0** | 0.505 | 0.176 | 1169.212 | 2.863 | 0.081 | |
| **A0T0 - A2T1** | 3.503 | 0.508 | 1187.817 | 6.899 | <0.001 | *** |
| **A0T0 - A2T2** | 3.204 | 0.413 | 1166.479 | 7.762 | <0.001 | *** |
| **A0T0 - A2T3** | 3.327 | 0.572 | 1245.098 | 5.819 | <0.001 | *** |
| **A0T0 - A2T4** | 5.087 | 0.333 | 1204.6 | 15.253 | <0.001 | *** |
| **A0T0 - Atypical** | 1.503 | 0.19 | 1160.247 | 7.914 | <0.001 | *** |
| **A1T0 - A2T0** | 0.128 | 0.193 | 1155.312 | 0.663 | 0.998 | |
| **A1T0 - A2T1** | 3.126 | 0.513 | 1183.802 | 6.089 | <0.001 | *** |
| **A1T0 - A2T2** | 2.827 | 0.42 | 1163.178 | 6.729 | <0.001 | *** |
| **A1T0 - A2T3** | 2.95 | 0.577 | 1241.252 | 5.112 | <0.001 | *** |
| **A1T0 - A2T4** | 4.71 | 0.343 | 1199.901 | 13.725 | <0.001 | *** |
| **A1T0 - Atypical** | 1.127 | 0.206 | 1153.067 | 5.457 | <0.001 | *** |
| **A2T0 - A2T1** | 2.998 | 0.512 | 1178.576 | 5.855 | <0.001 | *** |
| **A2T0 - A2T2** | 2.699 | 0.419 | 1156.606 | 6.446 | <0.001 | *** |
| **A2T0 - A2T3** | 2.822 | 0.576 | 1238.442 | 4.895 | <0.001 | *** |
| **A2T0 - A2T4** | 4.582 | 0.342 | 1194.454 | 13.381 | <0.001 | *** |
| **A2T0 - Atypical** | 0.998 | 0.206 | 1145.364 | 4.846 | <0.001 | *** |
| **A2T1 - A2T2** | -0.299 | 0.632 | 1169.789 | -0.473 | 1 | |
| **A2T1 - A2T3** | -0.176 | 0.746 | 1214.924 | -0.236 | 1 | |
| **A2T1 - A2T4** | 1.584 | 0.585 | 1188.357 | 2.706 | 0.122 | |
| **A2T1 - Atypical** | -2 | 0.518 | 1179.987 | -3.863 | 0.003 | ** |
| **A2T2 - A2T3** | 0.123 | 0.686 | 1213.78 | 0.179 | 1 | |
| **A2T2 - A2T4** | 1.883 | 0.505 | 1174.795 | 3.727 | 0.005 | ** |
| **A2T2 - Atypical** | -1.701 | 0.425 | 1157.849 | -3.999 | 0.002 | ** |
| **A2T3 - A2T4** | 1.76 | 0.642 | 1234.916 | 2.74 | 0.112 | |
| **A2T3 - Atypical** | -1.824 | 0.581 | 1237.892 | -3.137 | 0.037 | * |
| **A2T4 - Atypical** | -3.583 | 0.35 | 1192.073 | -10.242 | <0.001 | *** |

**Supplementary Table 7. Post-hoc testing comparing estimated marginal means of the mixed effect model predicting longitudinal MMSE scores in the training set.**

| Contrast | Difference | SE | DF | T ratio | p-value | Annotation |
|---|---|---|---|---|---|---|
| **A0T0 - A1T0** | -0.21 | 0.266 | 602.126 | -0.789 | 0.994 | |
| **A0T0 - A2T0** | -1.02 | 0.195 | 619.397 | -5.225 | <0.001 | *** |
| **A0T0 - A2T1** | -1.991 | 0.524 | 602.918 | -3.802 | 0.004 | ** |
| **A0T0 - A2T2** | -5.25 | 0.765 | 613.674 | -6.866 | <0.001 | *** |
| **A0T0 - A2T3** | -5.848 | 0.493 | 621.746 | -11.868 | <0.001 | *** |
| **A0T0 - A2T4** | -5.696 | 0.324 | 643.266 | -17.56 | <0.001 | *** |
| **A0T0 - Atypical** | -2.269 | 0.25 | 623.257 | -9.079 | <0.001 | *** |
| **A1T0 - A2T0** | -0.811 | 0.291 | 606.155 | -2.79 | 0.099 | |
| **A1T0 - A2T1** | -1.782 | 0.566 | 602.08 | -3.147 | 0.037 | * |
| **A1T0 - A2T2** | -5.04 | 0.794 | 611.934 | -6.347 | <0.001 | *** |
| **A1T0 - A2T3** | -5.638 | 0.538 | 617.33 | -10.487 | <0.001 | *** |
| **A1T0 - A2T4** | -5.486 | 0.39 | 630.256 | -14.078 | <0.001 | *** |
| **A1T0 - Atypical** | -2.059 | 0.33 | 612.817 | -6.239 | <0.001 | *** |
| **A2T0 - A2T1** | -0.971 | 0.535 | 600.774 | -1.813 | 0.611 | |
| **A2T0 - A2T2** | -4.23 | 0.773 | 612.887 | -5.472 | <0.001 | *** |
| **A2T0 - A2T3** | -4.828 | 0.506 | 621.033 | -9.538 | <0.001 | *** |
| **A2T0 - A2T4** | -4.676 | 0.346 | 647.037 | -13.509 | <0.001 | *** |
| **A2T0 - Atypical** | -1.249 | 0.276 | 622.977 | -4.526 | <0.001 | *** |
| **A2T1 - A2T2** | -3.258 | 0.913 | 609.519 | -3.569 | 0.009 | ** |
| **A2T1 - A2T3** | -3.857 | 0.702 | 611.273 | -5.496 | <0.001 | *** |
| **A2T1 - A2T4** | -3.705 | 0.597 | 616.641 | -6.206 | <0.001 | *** |
| **A2T1 - Atypical** | -0.278 | 0.559 | 605.517 | -0.497 | 1 | |
| **A2T2 - A2T3** | -0.598 | 0.896 | 615.233 | -0.668 | 0.998 | |
| **A2T2 - A2T4** | -0.446 | 0.816 | 618.648 | -0.547 | 0.999 | |
| **A2T2 - Atypical** | 2.981 | 0.789 | 614.324 | 3.778 | 0.004 | ** |
| **A2T3 - A2T4** | 0.152 | 0.569 | 629.996 | 0.267 | 1 | |
| **A2T3 - Atypical** | 3.579 | 0.53 | 622.798 | 6.753 | <0.001 | *** |
| **A2T4 - Atypical** | 3.427 | 0.379 | 642.44 | 9.036 | <0.001 | *** |

**Supplementary Table 8. Post-hoc testing comparing estimated marginal means of the mixed effect model predicting longitudinal CDR-SB scores in the validation set.**

| Contrast | Difference | SE | DF | T ratio | p-value | Annotation |
|---|---|---|---|---|---|---|
| **A0T0 - A1T0** | 0.122 | 0.399 | 435.833 | 0.306 | 1 | |
| **A0T0 - A2T0** | 0.765 | 0.307 | 443.616 | 2.493 | 0.201 | |
| **A0T0 - A2T1** | 1.844 | 0.794 | 434.019 | 2.324 | 0.283 | |
| **A0T0 - A2T2** | 5.782 | 1.283 | 495.745 | 4.507 | <0.001 | *** |
| **A0T0 - A2T3** | 6.925 | 1.014 | 433.821 | 6.828 | <0.001 | *** |
| **A0T0 - A2T4** | 7.719 | 0.626 | 457.473 | 12.338 | <0.001 | *** |
| **A0T0 - Atypical** | 3.651 | 0.426 | 455.481 | 8.577 | <0.001 | *** |
| **A1T0 - A2T0** | 0.642 | 0.45 | 439.859 | 1.428 | 0.844 | |
| **A1T0 - A2T1** | 1.722 | 0.86 | 434.874 | 2.003 | 0.481 | |
| **A1T0 - A2T2** | 5.66 | 1.324 | 491.599 | 4.275 | 0.001 | *** |
| **A1T0 - A2T3** | 6.803 | 1.066 | 433.669 | 6.383 | <0.001 | *** |
| **A1T0 - A2T4** | 7.597 | 0.707 | 452.516 | 10.751 | <0.001 | *** |
| **A1T0 - Atypical** | 3.529 | 0.538 | 448.188 | 6.56 | <0.001 | *** |
| **A2T0 - A2T1** | 1.079 | 0.816 | 432.288 | 1.323 | 0.89 | |
| **A2T0 - A2T2** | 5.018 | 1.296 | 493.055 | 3.871 | 0.003 | ** |
| **A2T0 - A2T3** | 6.161 | 1.032 | 432.622 | 5.968 | <0.001 | *** |
| **A2T0 - A2T4** | 6.954 | 0.66 | 458.315 | 10.535 | <0.001 | *** |
| **A2T0 - Atypical** | 2.887 | 0.47 | 450.432 | 6.144 | <0.001 | *** |
| **A2T1 - A2T2** | 3.938 | 1.489 | 478.407 | 2.644 | 0.143 | |
| **A2T1 - A2T3** | 5.081 | 1.267 | 433.902 | 4.009 | 0.002 | ** |
| **A2T1 - A2T4** | 5.875 | 0.988 | 445.393 | 5.948 | <0.001 | *** |
| **A2T1 - Atypical** | 1.807 | 0.869 | 437.554 | 2.079 | 0.43 | |
| **A2T2 - A2T3** | 1.143 | 1.615 | 469.264 | 0.708 | 0.997 | |
| **A2T2 - A2T4** | 1.937 | 1.409 | 489.511 | 1.375 | 0.869 | |
| **A2T2 - Atypical** | -2.131 | 1.33 | 492.294 | -1.602 | 0.749 | |
| **A2T3 - A2T4** | 0.794 | 1.169 | 439.739 | 0.679 | 0.997 | |
| **A2T3 - Atypical** | -3.274 | 1.074 | 435.705 | -3.048 | 0.05 | * |
| **A2T4 - Atypical** | -4.068 | 0.723 | 459.506 | -5.63 | <0.001 | *** |

**Supplementary Table 9. Post-hoc testing comparing estimated marginal means of the mixed effect model predicting longitudinal MMSE scores in the validation set.**

| Contrast | Log-rank statistic | p-value | Annotation |
|---|---|---|---|
| **A0T0-A1T0** | 3.59 | 0.058 | |
| **A0T0-A2T0** | 7.04 | 0.008 | ** |
| **A0T0-A2T1** | 49.52 | <0.001 | *** |
| **A0T0-A2T2** | 81.78 | <0.001 | *** |
| **A0T0-A2T3** | 97.21 | <0.001 | *** |
| **A0T0-A2T4** | 197.65 | <0.001 | *** |
| **A1T0-A2T0** | 0.09 | 0.767 | |
| **A1T0-A2T1** | 16.31 | <0.001 | *** |
| **A1T0-A2T2** | 30.43 | <0.001 | *** |
| **A1T0-A2T3** | 41.33 | <0.001 | *** |
| **A1T0-A2T4** | 81.34 | <0.001 | *** |
| **A2T0-A2T1** | 16.1 | <0.001 | *** |
| **A2T0-A2T2** | 29.75 | <0.001 | *** |
| **A2T0-A2T3** | 31.83 | <0.001 | *** |
| **A2T0-A2T4** | 85.14 | <0.001 | *** |
| **A2T1-A2T2** | 0.08 | 0.78 | |
| **A2T1-A2T3** | 1.13 | 0.287 | |
| **A2T1-A2T4** | 1.84 | 0.175 | |
| **A2T2-A2T3** | 1.73 | 0.189 | |
| **A2T2-A2T4** | 2.79 | 0.095 | |
| **A2T3-A2T4** | 0.03 | 0.861 | |

**Supplementary Table 10. Results of pairwise log-rank tests comparing survival curves (CDR<1) in the training data.**

| Contrast | Log-rank statistic | p-value | Annotation |
|---|---|---|---|
| **A0T0-A1T0** | 0 | 1 | |
| **A0T0-A2T0** | 28.82 | <0.001 | *** |
| **A0T0-A2T1** | 15 | <0.001 | *** |
| **A0T0-A2T2** | 216.4 | <0.001 | *** |
| **A0T0-A2T3** | 143.85 | <0.001 | *** |
| **A0T0-A2T4** | 223.76 | <0.001 | *** |
| **A1T0-A2T0** | 5.88 | 0.015 | * |
| **A1T0-A2T1** | 2 | 0.157 | |
| **A1T0-A2T2** | 45.54 | <0.001 | *** |
| **A1T0-A2T3** | 29.49 | <0.001 | *** |
| **A1T0-A2T4** | 44.73 | <0.001 | *** |
| **A2T0-A2T1** | 0.01 | 0.935 | |
| **A2T0-A2T2** | 29.33 | <0.001 | *** |
| **A2T0-A2T3** | 17.79 | <0.001 | *** |
| **A2T0-A2T4** | 43.8 | <0.001 | *** |
| **A2T1-A2T2** | 7.5 | 0.006 | ** |
| **A2T1-A2T3** | 4.99 | 0.025 | * |
| **A2T1-A2T4** | 5.6 | 0.018 | * |
| **A2T2-A2T3** | 1.12 | 0.29 | |
| **A2T2-A2T4** | 0.3 | 0.583 | |
| **A2T3-A2T4** | 0.57 | 0.451 | |

**Supplementary Table 11. Results of pairwise log-rank tests comparing survival curves (CDR<1) in the validation data.**

| Variable | NS | A1T0-A2T0 | A2T1-A2T4 | p | A1T0-A2T0 vs. NS | A2T1-A2T4 vs. NS |
|---|---|---|---|---|---|---|
| **Age** | 73.616 | 73.122 | 75.539 | <0.001 | 0.899 | 0.133 |
| **Sex (Male)** | 45.8 (82/179) | 40.2 (206/513) | 52.6 (72/137) | 0.026 | 0.362 | 0.383 |
| **Education** | 16.098 | 16.43 | 15.898 | <0.001 | 0.383 | 0.743 |
| **BMI** | 26.913 | 27.524 | 26.881 | 0.31 | NA | NA |
| **APOE E4 (carrier)** | 58.1 (104/179) | 62 (318/513) | 66.4 (91/137) | 0.067 | NA | NA |
| **MMSE** | 27.739 | 28.602 | 25.91 | <0.001 | 0.002** | <0.001*** |
| **CDR-SB** | 0.88 | 0.387 | 2.319 | <0.001 | 0.016* | <0.001*** |
| **PAC-Frontal** | 0.032 | 0.032 | -0.16 | <0.001 | 0.989 | 0.197 |
| **PAC-Parietal** | -0.098 | 0.003 | 0.116 | 0.19 | NA | NA |
| **PAC-Occipital** | -0.042 | -0.09 | 0.39 | <0.001 | 0.989 | 0.167 |
| **PAC-Sensorimotor** | -0.065 | 0.104 | -0.306 | <0.001 | 0.251 | 0.197 |
| **PTC-LeftParietalTemporal** | 0.151 | 0.008 | -0.227 | <0.001 | 0.758 | 0.251 |
| **PTC-Occipital** | 0.334 | -0.078 | -0.142 | <0.001 | 0.278 | 0.383 |
| **PTC-RightParietalTemporal** | 0.137 | -0.119 | 0.266 | <0.001 | 0.383 | 0.743 |
| **PTC-MedialTemporal** | 0.246 | -0.162 | 0.286 | <0.001 | 0.001** | 0.989 |
| **PTC-Sensorimotor** | 0.33 | -0.124 | 0.031 | <0.001 | 0.012* | 0.383 |
| **PTC-Frontal** | 0.312 | -0.091 | -0.068 | <0.001 | 0.096 | 0.362 |
| **PTC-InsularMedialFrontal** | 0.349 | -0.063 | -0.219 | <0.001 | 0.012* | 0.009** |

**Supplementary Table 12. Models comparing non-stageable (NS) individuals against stageable individuals with only amyloid pathology (A1T0-A2T0) or with amyloid and tau pathology (A2T1-A2T4) in training-ADS.** Columns 2-4 show the mean (continuous variables) or the percentage and fraction (categorical variables) for each group. Continuous values were compared with a one-way ANCOVAs adjusted for age and sex (except when used as the dependent variable), while categorical variables were compared with chi-squared tests. The p column shows the p-value for the overall model, while the final two columns show post-hoc p-values for comparisons between groups. Post-hoc comparisons were not performed for non-significant models, and are marked with NA. The Variables beginning with "PAC-" and "PTC-" are W-scores in PACs and PTCs, and they are adjusted for the global amyloid and tau burden (respectively).

| Variable | NS | A1T0-A2T0 | A2T1-A2T4 | p | A1T0-A2T0 vs. NS | A2T1-A2T4 vs. NS |
|---|---|---|---|---|---|---|
| **Age** | 71.925 | 73.644 | 71.499 | <0.001 | 0.307 | 0.98 |
| **Sex (Male)** | 42.1 (51/121) | 42.2 (181/429) | 42.4 (75/177) | 0.999 | NA | NA |
| **Education** | 15.508 | 15.723 | 15.989 | <0.001 | 0.98 | 0.801 |
| **BMI** | 29.537 | 28.59 | 27.204 | <0.001 | 0.98 | 0.307 |
| **APOE E4 (carrier)** | 66.9 (81/121) | 67.1 (288/429) | 59.3 (105/177) | 0.211 | NA | NA |
| **MMSE** | 26.316 | 28.178 | 25 | <0.001 | <0.001*** | 0.096 |
| **CDR-SB** | 1.799 | 0.624 | 3.821 | <0.001 | <0.001*** | <0.001*** |
| **PAC-Frontal** | -0.05 | 0.075 | -0.147 | 0.03 | 0.98 | 0.98 |
| **PAC-Parietal** | -0.032 | -0.005 | 0.035 | 0.019 | 0.98 | 0.98 |
| **PAC-Occipital** | -0.213 | -0.044 | 0.252 | 0.161 | NA | NA |
| **PAC-Sensorimotor** | -0.118 | 0.108 | -0.181 | 0.002 | 0.801 | 0.98 |
| **PTC-LeftParietalTemporal** | -0.082 | -0.001 | 0.059 | <0.001 | 0.98 | 0.98 |
| **PTC-Occipital** | -0.022 | -0.093 | 0.241 | <0.001 | 0.98 | 0.98 |
| **PTC-RightParietalTemporal** | -0.132 | 0.094 | -0.138 | <0.001 | 0.519 | 0.98 |
| **PTC-MedialTemporal** | 0.367 | -0.216 | 0.272 | <0.001 | 0.002** | 0.98 |
| **PTC-Sensorimotor** | -0.019 | 0.019 | -0.032 | <0.001 | 0.98 | 0.98 |
| **PTC-Frontal** | -0.04 | 0.083 | -0.173 | <0.001 | 0.98 | 0.98 |
| **PTC-InsularMedialFrontal** | 0.162 | 0.017 | -0.152 | <0.001 | 0.98 | 0.55 |

**Supplementary Table 13. Models comparing non-stageable (NS) individuals against stageable individuals with only amyloid pathology (A1T0-A2T0) or with amyloid and tau pathology (A2T1-A2T4) in validation-ADS.** Columns 2-4 show the mean (continuous variables) or the percentage and fraction (categorical variables) for each group. Continuous values were compared with a one-way ANCOVAs adjusted for age and sex (except when used as the dependent variable), while categorical variables were compared with chi-squared tests. The p column shows the p-value for the overall model, while the final two columns show post-hoc p-values for comparisons between groups. Post-hoc comparisons were not performed for non-significant models, and are marked with NA. The Variables beginning with "PAC-" and "PTC-" are W-scores in PACs and PTCs, and they are adjusted for the global amyloid and tau burden (respectively).

| Variable | Expected | Resilient | Vulnerable | p | Resilient vs. Expected | Vulnerable vs. Expected |
|---|---|---|---|---|---|---|
| **Age** | 72.989 | 73.713 | 75.612 | <0.001 | 0.999 | 0.002** |
| **Sex (Male)** | 36.9 (100/271) | 52.7 (29/55) | 49.5 (103/208) | 0.008 | 0.075 | 0.017* |
| **Education** | 16.561 | 16.036 | 15.938 | <0.001 | 0.246 | 0.012* |
| **BMI** | 27.652 | 27.08 | 27.828 | 0.201 | NA | NA |
| **APOE E4 (carrier)** | 52 (141/271) | 69.1 (38/55) | 54.3 (113/208) | 0.02 | 0.019* | 0.999 |
| **MMSE** | 28.466 | 26.889 | 27.338 | <0.001 | 0.002** | 0.007** |
| **CDR-SB** | 0.6 | 0.9 | 0.834 | <0.001 | 0.938 | 0.999 |
| **Gray matter volume** | 0.378 | 0.36 | 0.372 | <0.001 | <0.001*** | 0.999 |
| **Amyloid SUVR** | 1.48 | 1.673 | 1.486 | <0.001 | <0.001*** | 0.783 |
| **Tau SUVR** | 1.221 | 1.571 | 1.167 | <0.001 | <0.001*** | 0.015* |

**Supplementary Table 14. Models comparing expected, resilient, and vulnerable individuals in training-ADS.** Columns 2-4 show the mean (continuous variables) or the percentage and fraction (categorical variables) for each group. Continuous values were compared with a one-way ANCOVAs adjusted for age and sex (except when used as the dependent variable), while categorical variables were compared with chi-squared tests. The p column shows the p-value for the overall model, while the final two columns show post-hoc p-values for comparisons between groups. Post-hoc comparisons were not performed for non-significant models, and are marked with NA.

| Variable | Expected | Resilient | Vulnerable | p | Resilient vs. Expected | Vulnerable vs. Expected |
|---|---|---|---|---|---|---|
| **Age** | 71.701 | 71.829 | 75.344 | 0.001 | 1 | 0.002** |
| **Sex (Male)** | 41.2 (114/277) | 35.5 (11/31) | 36.9 (31/84) | 0.688 | NA | NA |
| **Education** | 15.754 | 15.645 | 14.571 | <0.001 | 1 | <0.001*** |
| **BMI** | 28.487 | 32.22 | 30.463 | 0.001 | 0.21 | 0.124 |
| **APOE E4 (carrier)** | 75.5 (209/277) | 77.4 (24/31) | 71.4 (60/84) | 0.603 | NA | NA |
| **MMSE** | 28.13 | 26.867 | 25.051 | <0.001 | 0.159 | <0.001*** |
| **CDR-SB** | 1.403 | 1.177 | 2.464 | 0.008 | 1 | 0.015* |
| **Gray matter volume** | 0.371 | 0.367 | 0.368 | <0.001 | 0.67 | 1 |
| **Amyloid SUVR** | 1.606 | 1.707 | 1.576 | 0.014 | 0.318 | 0.67 |
| **Tau SUVR** | 1.277 | 1.569 | 1.194 | <0.001 | <0.001*** | 0.365 |

**Supplementary Table 15. Models comparing expected, resilient, and vulnerable individuals in validation-ADS.** Columns 2-4 show the mean (continuous variables) or the percentage and fraction (categorical variables) for each group. Continuous values were compared with a one-way ANCOVAs adjusted for age and sex (except when used as the dependent variable), while categorical variables were compared with chi-squared tests. The p column shows the p-value for the overall model, while the final two columns show post-hoc p-values for comparisons between groups. Post-hoc comparisons were not performed for non-significant models, and are marked with NA.
